# PanEcho: Complete AI-enabled echocardiography interpretation with multi-task deep learning

**DOI:** 10.1101/2024.11.16.24317431

**Authors:** Gregory Holste, Evangelos K. Oikonomou, Márton Tokodi, Attila Kovács, Zhangyang Wang, Rohan Khera

**Affiliations:** Department of Electrical and Computer Engineering, The University of Texas at Austin, Austin, TX, USA; Section of Cardiovascular Medicine, Department of Internal Medicine, Yale School of Medicine, New Haven, CT, USA; Cardiovascular Data Science (CarDS) Lab, Yale School of Medicine, New Haven, CT, USA; Department of Experimental Cardiology and Surgical Techniques, Heart and Vascular Center, Semmelweis University, Budapest, Hungary; Institute for Clinical Data Management, Semmelweis University, Budapest, Hungary; Section of Health Informatics, Department of Biostatistics, Yale School of Public Health, New Haven, CT, USA; Center for Outcomes Research and Evaluation, Yale-New Haven Hospital, New Haven, CT, USA; Section of Biomedical Informatics and Data Science, Yale School of Medicine, New Haven, CT, USA

## Abstract

**Importance:** Echocardiography is a cornerstone of cardiovascular care but relies on expert interpretation and manual reporting from a series of videos. We propose an artificial intelligence (AI) system, PanEcho, to automate echocardiogram interpretation with multi-task deep learning.

**Objective:** To develop and evaluate the accuracy of PanEcho on a comprehensive set of 39 echocardiographic labels and measurements on transthoracic echocardiography (TTE).

**Design, Setting, and Participants:** This study represents the development and retrospective, multi-site validation of an AI system. PanEcho was developed using a sample of TTE studies conducted at Yale-New Haven Health System (YNHHS) hospitals and clinics from January 2016-June 2022 during routine care. The trained model was internally validated in a temporally distinct YNHHS cohort from July-December 2022, externally validated across four diverse external cohorts, and made publicly available.

**Main Outcomes and Measures:** The primary outcome was the area under the receiver operating characteristic curve (AUC) for diagnostic classification tasks and mean absolute error (MAE) for parameter estimation tasks, comparing AI predictions with the assessment of the interpreting cardiologist.

**Results:** This study included 1.2 million echocardiographic videos from 32,265 TTE studies of 24,405 patients across YNHHS hospitals and clinics. PanEcho performed 18 diagnostic classification tasks with a median AUC of 0.91 (IQR: 0.88-0.93) and estimated 21 echocardiographic parameters with a median normalized MAE of 0.13 (0.10-0.18) in internal validation. For instance, the model accurately estimated left ventricular (LV) ejection fraction (MAE: 4.2% internal; 4.5% external) and detected moderate or higher LV systolic dysfunction (AUC: 0.98 internal; 0.99 external), RV systolic dysfunction (0.93 internal; 0.94 external), and severe aortic stenosis (0.98 internal; 1.00 external). PanEcho maintained excellent performance in limited imaging protocols, performing 15 diagnosis tasks with 0.91 median AUC (IQR: 0.87-0.94) in an abbreviated TTE cohort and 14 tasks with 0.85 median AUC (0.77-0.87) on real-world point-of-care ultrasound acquisitions by non-experts from YNHHS emergency departments.

**Conclusions and Relevance:** We report an AI system that automatically interprets echocardiograms, maintaining high accuracy across geography and time from complete and limited studies. PanEcho may be used as an adjunct reader in echocardiography labs or rapid AI-enabled screening tool in point-of-care settings.

**KEY POINTS:** *Question:* Can artificial intelligence (AI) fully automate echocardiogram interpretation?

*Findings:* We report the development and validation of an automated AI system for echocardiogram analysis, called *PanEcho*, that performed 18 diagnostic classification tasks with a median area under the receiver operating characteristic curve (AUC) of 0.91 and 21 echocardiographic parameter estimation tasks with a median normalized mean absolute error (MAE) of 0.14.

*Meaning:* An AI system can automate complete echocardiogram interpretation with high accuracy, potentially accelerating workflows and enabling rapid cardiovascular health screening in point-of-care settings with limited access to trained experts.

## INTRODUCTION

Echocardiography is a pillar of cardiovascular diagnostics, providing in-depth phenotyping of cardiac, valvular, and vascular structure and function.^1^ More than 7.5 million echocardiographic studies are performed annually in the United States alone, and increasing referrals contribute to rising healthcare expenditures across most nations.^2,3^ Accurate reporting of echocardiography requires time, skilled acquisition, and expert readers but remains limited by the availability of sufficient experts and substantial inter-rater variability.^4,5^ Reliance on scarcely available expert interpretation poses a barrier to access to this important diagnostic modality, especially given the increasing accessibility of handheld ultrasound devices at the point of care.^6,7^

Artificial intelligence (AI) algorithms have shown promise in automating various aspects of echocardiography reporting, from detecting valvular abnormalities^8–11^ to quantifying key measurements such as left ventricular (LV) ejection fraction (EF),^12–17^ and others.^18–23^ However, existing solutions typically rely on single-view inputs and are limited to single tasks.^8,11,13,20,22–26^ This process differs from clinical practice, where multiple views and imaging modes, such as color Doppler imaging, are integrated to form a comprehensive evaluation, spanning functional and structural metrics of all major chambers, valves, and vessels. Versatile AI systems that handle this multi-view, multi-task workflow accurately and robustly would enable efficient, reader-independent phenotyping of echocardiography but are currently lacking.

To bridge this gap and provide a scalable solution for fully automated echocardiographic interpretation, we sought to develop and validate an end-to-end, view-agnostic deep learning model capable of simultaneously performing the full range of key echocardiographic reporting tasks. We present the development of PanEcho, a novel AI model that uses multi-task deep learning on over one million standard 2D B-mode and color Doppler echocardiogram videos to perform 39 diverse interpretation tasks and report its internal and external validation across a range of acquisition protocols. In addition to this study report, we have publicly released the model and source code to accelerate research on AI-enabled echocardiographic interpretation (https://github.com/CarDS-Yale/PanEcho).

## METHODS

### Data Source and Patient Population

This study was approved by the Yale University and local Institutional Review Boards, and the need for informed consent was waived since this research represents secondary analysis of existing data.

#### PanEcho development and internal validation

Data for model development and internal validation was derived from transthoracic echocardiography (TTE) studies performed at Yale-New Haven Health System (YNHHS) hospitals from 2016-2022 during routine clinical care. We used our previously published approach^8^ to extract and deidentify echocardiographic videos by masking out pixels containing protected health information. This study included 2D grayscale and color Doppler videos from all echocardiographic views. The YNHHS cohort was split into development and validation sets, with studies from July to December 2022 set aside as a temporally distinct internal validation set. The remaining studies from January 2016 to June 2022 were used for model development after removing studies from all validation set patients to prevent data leakage. The development set was randomly partitioned into training (92.5%) and tuning (7.5%) sets at the patient level for model training. See the **eMethods** for full details on YNHHS dataset curation and preprocessing.

#### External validation of PanEcho

This study featured four cohorts for external validation: RVENet+, POCUS, EchoNet-Dynamic, and EchoNet-LVH, representing a broad range of phenotypes with standard and point-of-care echocardiographic acquisitions across time and geography. The RVENet+ cohort consisted of complete echocardiographic studies conducted at the Heart and Vascular Center of Semmelweis University in Budapest, Hungary from 2013 to 2021 and included all available 2D views. As previously described,^27,28^ this cohort comprised seven distinct subpopulations ranging from elite athletes to heart transplant recipients, representing a diverse population. This cohort underwent the same deidentification and preprocessing steps as the YNHHS cohort.

The POCUS cohort consisted of consecutive patients who underwent cardiac-focused POCUS imaging as part of care in YNHHS-affiliated emergency departments (EDs) from 2016-2023. In contrast to all other cohorts that included studies performed by echocardiography technicians, these abbreviated scans were performed by ED providers. We present analysis in a subset of patients with POCUS imaging and a temporally linked complete echocardiographic exam performed no more than a day apart, essential to provide reliable ground truth labels. This cohort underwent the same deidentification and preprocessing steps as the YNHHS and RVENet+ cohorts, ensuring no patient overlap with the YNHHS cohort. Additional quality control was needed to ensure minimal diagnostic quality in these noisy acquisitions by non-experts, removing non-cardiac views and filtering for studies with at least three of these five key views: parasternal long axis (PLAX), parasternal short axis (PSAX) at the papillary level, A4C, apical 2-chamber (A2C), and apical 5-chamber (A5C). Full POCUS cohort curation details are described in previous work^29^ and the **eMethods**. See **Figure 1A** for an exclusion flowchart of all key internal and external cohorts.

**Figure 1.**
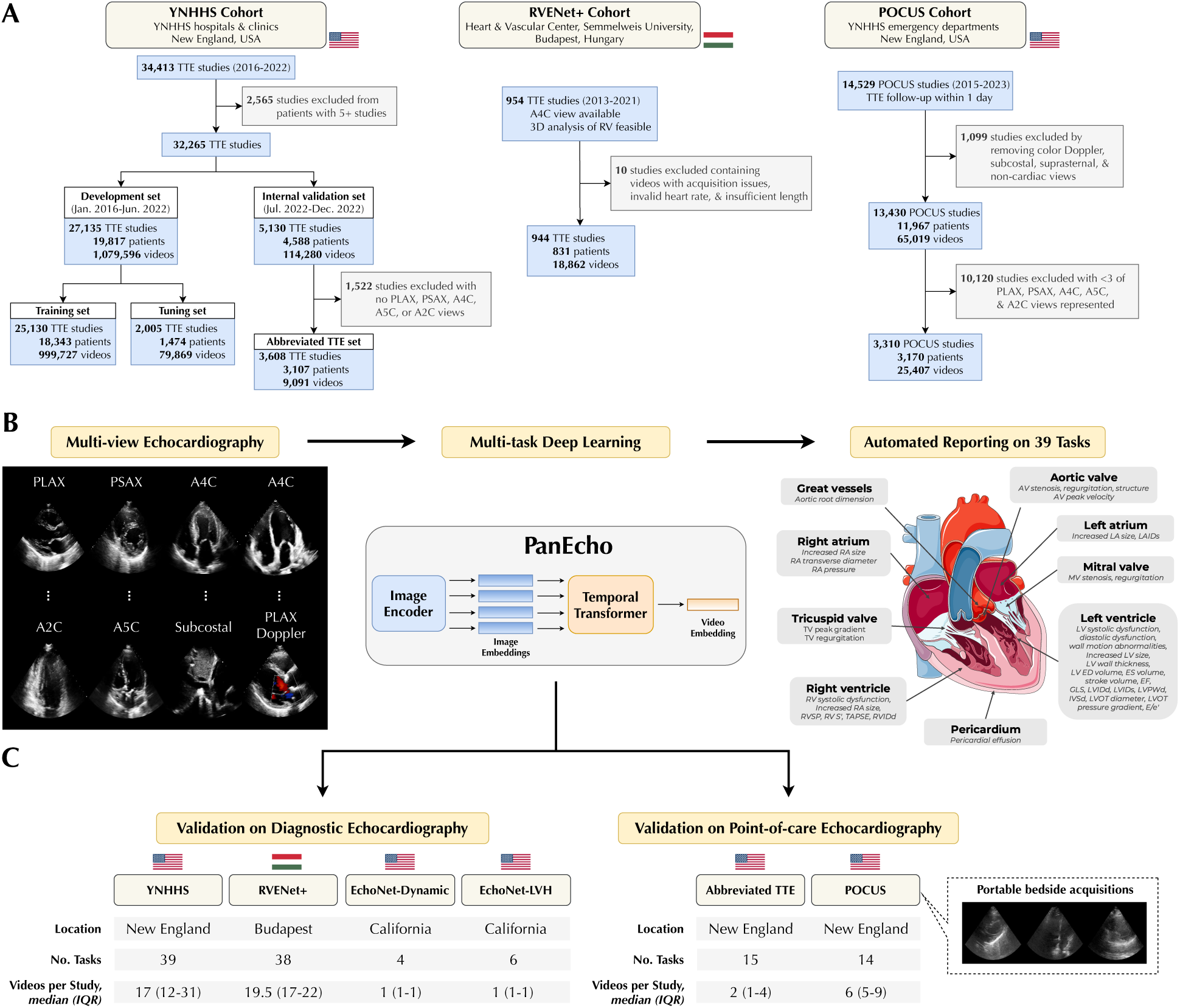
PanEcho Model Development and Study Design. Diagram of PanEcho workflow and study design with flowchart describing internal and external validation cohorts (A). PanEcho is an AI model that analyzes all 2D views acquired during a transthoracic echocardiogram and automatically performs 39 key echocardiographic interpretation tasks (B). The model is externally validated on complete diagnostic-quality echocardiograms as well as limited acquisitions acquired at the point of care (C). A2C = apical 2-chamber; A4C = apical 4-chamber; A5C = apical 5-chamber; AV = aortic valve; ED = end-diastolic; ES = end-systolic; IVSd = interventricular septum thickness diastole; LA = left atrium; LV = left ventricle; LAIDs = left atrial internal diameter systole; LVIDd = left ventricular internal diameter diastole; LVIDs = left ventricular internal diameter systole; LVOT = left ventricular outflow tract; LVPWd = left ventricular posterior wall thickness diastole; PLAX = parasternal long axis; POCUS = point-of-care ultrasound; PSAX = parasternal short axis; PG = pressure gradient; RA = right atrium; RV = right ventricle; RVIDd = right ventricular internal diameter diastole; RV S’ = right ventricular systolic excursion velocity; TTE = transthoracic echocardiography; TV = tricuspid valve; YNHHS = Yale-New Haven Health System.

EchoNet-Dynamic^13^ and EchoNet-LVH^24^ are publicly available datasets for assessment of LV function and structure, respectively, that were included as additional external validation cohorts in this study. EchoNet-Dynamic consists of 10,030 individual A4C videos from unique echocardiograms performed at Stanford University Hospital from 2016-2018 with measurements of LV function and volumes, and EchoNet-LVH consists of 12,000 PLAX videos from echocardiograms performed at Stanford Health Care from 2008-2020 with measurements of LV dimensions.

### Echocardiographic Reporting Labels

For each study in the YNHHS, RVENet+, and POCUS cohorts, the team of investigators jointly reviewed and defined a list of 39 key reporting tasks, representing a wide variety of categorical findings and continuous measurements covering all major axes of echocardiographic reporting. This included 18 classification tasks capturing the size, structure, and function of all four heart chambers, valvular disease, etc., and 21 regression tasks quantifying key dimensions of each chamber, blood flow velocities, and more. Labels were extracted from the local electronic echocardiography reporting systems and reflected the final measurements and reporting confirmed by a certified echocardiographer. All interpretations were performed in line with standard guidelines of the American Society of Echocardiography^1^ and to the discretion of the echocardiographer; descriptions of all diagnostic labels are detailed in **eTable 1**. Finally, we extracted a total of 10 labels from EchoNet-Dynamic and EchoNet-LVH based on the LV measurements provided, such as LVEF, LV interventricular septum thickness (IVSd), and others. Since categorical labels were not explicitly provided, we determined these via thresholds described in the **eMethods.**

### PanEcho Model Development

We designed a multi-task, video-based deep learning model with a 2D image encoder, temporal Transformer, and output heads dedicated to individual tasks (**Figure 1B**). In its design, PanEcho mimics a human reader, integrating all available echocardiographic views and assessing all 39 echocardiographic reporting tasks described above. Each video frame is first processed by the 2D image encoder, a convolutional neural network (CNN), which produces a learned embedding of each frame. These frame-wise embeddings are then interpreted as an ordered sequence and modeled using self-attention^30^ to learn time-varying associations over the frames. Finally, a video-level embedding is formed and used as input to the task-specific output heads. Rather than develop separate view- and task-specific models, this design efficiently shares computation across views with a unified view-agnostic encoder and parallelizes task-specific modeling with lightweight output heads specialized for each task. PanEcho was trained end-to-end to minimize the mean of all task-specific losses. See the **eMethods** for full implementation details.

### Multi-task Evaluation on Diagnostic Echocardiography

PanEcho was first evaluated across all 39 labels in the internal YNHHS validation set as well as the three external cohorts RVENet+ (38 labels), EchoNet-Dynamic (4 labels), and EchoNet-LVH (6 labels) with diagnostic-quality videos from complete echocardiographic studies (**Figure 1C**). Since interpretations are unique to each echocardiographic study, evaluation was performed at the study level using all available videos and tasks. For multi-view datasets like the YNHHS and RVENet+ cohorts, PanEcho automatically aggregated its predictions across all videos acquired during a study at inference time to form study-level interpretations. In contrast, EchoNet-Dynamic and EchoNet-LVH feature one video per study, evaluating PanEcho’s ability to provide interpretations from a single view.

### Multi-task Evaluation on Point-of-care Echocardiography

To illustrate the versatility of PanEcho across imaging protocols, we also evaluated its performance in both a simulated abbreviated TTE protocol and a real-world point-of-care setting. To simulate an abbreviated acquisition, inference was performed on the internal YNHHS validation set, but the model was only given access to a single video from each of the following key views per study if available: PLAX, PSAX at the papillary level, A4C, A5C, A2C. Validation was also performed across actual point-of-care acquisitions from YNHHS EDs in the POCUS cohort described above. Since POCUS imaging is focused on ruling out key abnormalities and lacks the protocolized approach for reliable estimation of linear or volumetric measurements, evaluation was restricted to all 15 classification tasks evaluable on grayscale 2D echocardiography; this excluded estimation tasks and diagnostic findings like valvular regurgitation, which require Doppler imaging to properly assess. See the **eMethods** for full details on point-of-care validation.

### Statistical Analysis

Categorical values are summarized as count (%), whereas continuous variables are summarized as median (IQR) to account for skewness. Classification tasks were primarily evaluated by area under the receiver operating characteristic curve (AUC) in addition to sensitivity, specificity, average precision, and Brier score; class-specific thresholds were determined by maximizing Youden’s index on the tuning set. Regression tasks were primarily evaluated by mean absolute error (MAE) in addition to Pearson’s correlation coefficient (r), median absolute deviation, root-mean-squared error, R^2^, and normalized MAE (divided by the mean ground truth measurement). For multi-class classification tasks, we present AUC results on the most severe class in the main text to simplify the presentation. When summarizing performance across regression tasks with different units, we report median normalized MAE to account for variations in scale across measurements. We computed 95% confidence intervals for all metrics with 1,000 bootstrap samples at the study level using the percentile method. Performance analysis was performed using Python version 3.10.8.

## RESULTS

### Study Cohorts and Model Development

The YNHHS cohort included 1,193,876 echocardiographic videos comprising all 2D views from 32,265 transthoracic echocardiography studies of 24,405 unique patients across YNHHS hospitals (median [IQR] age: 69.0 [58.0-79.0] years, 16,819 [52.1%] males, 5,884 [19.2%] reported non-White). PanEcho was developed using 999,727 videos from 25,130 studies of 18,343 YNHHS patients from January 2016-June 2022, while 5,130 studies from 4,588 distinct patients from July-December 2022 were used as a temporally distinct internal validation set. This set featured a broad range of echocardiographic phenotypes with a median (IQR) LVEF of 61.4% (57.0-65.0), 284 (5.8%) studies with moderate or higher LV systolic dysfunction, 86 (1.8%) severe aortic stenosis, 612 (12.2%) moderate or higher mitral regurgitation, among others (**Table 1**).

**Table 1.**
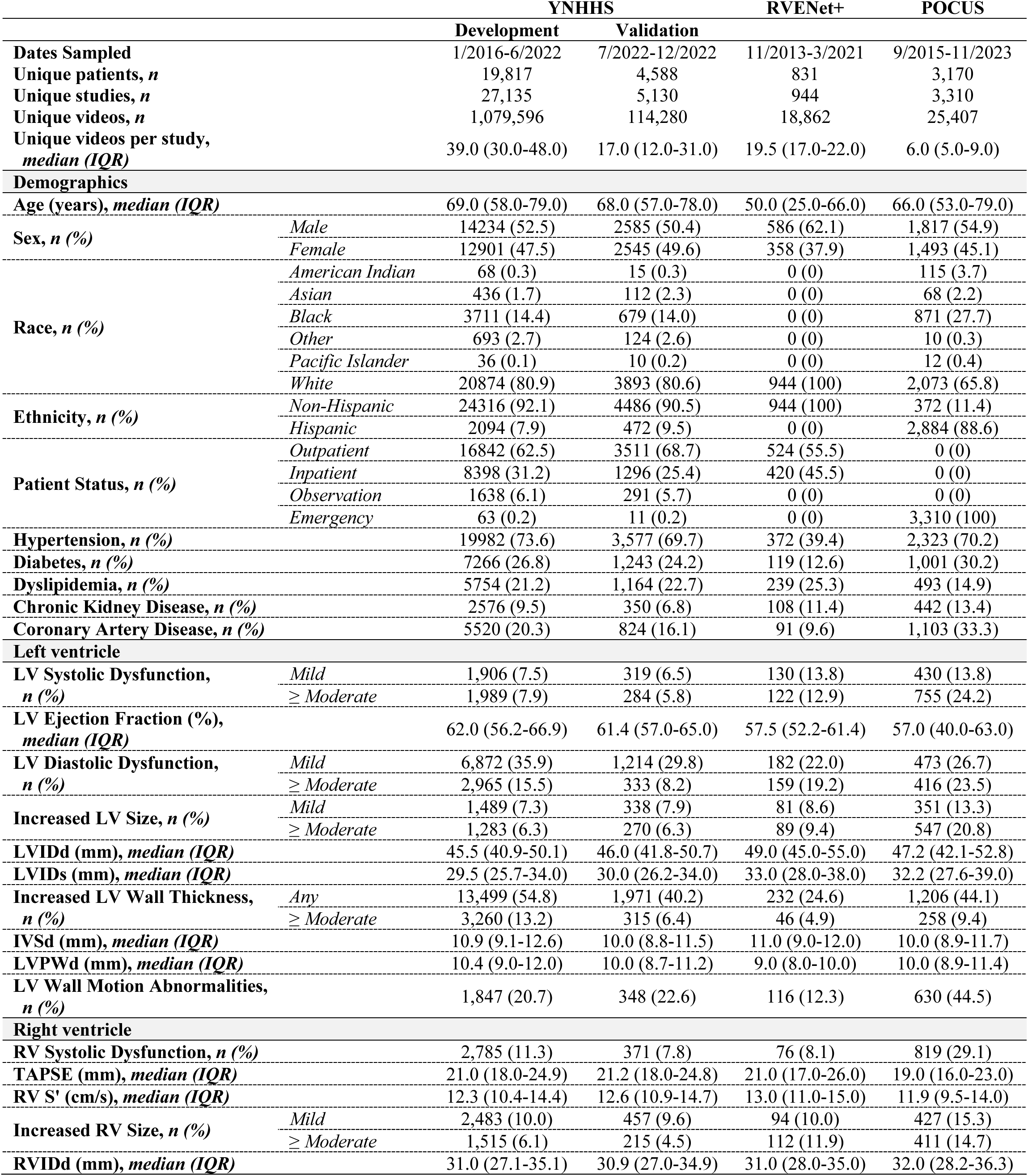
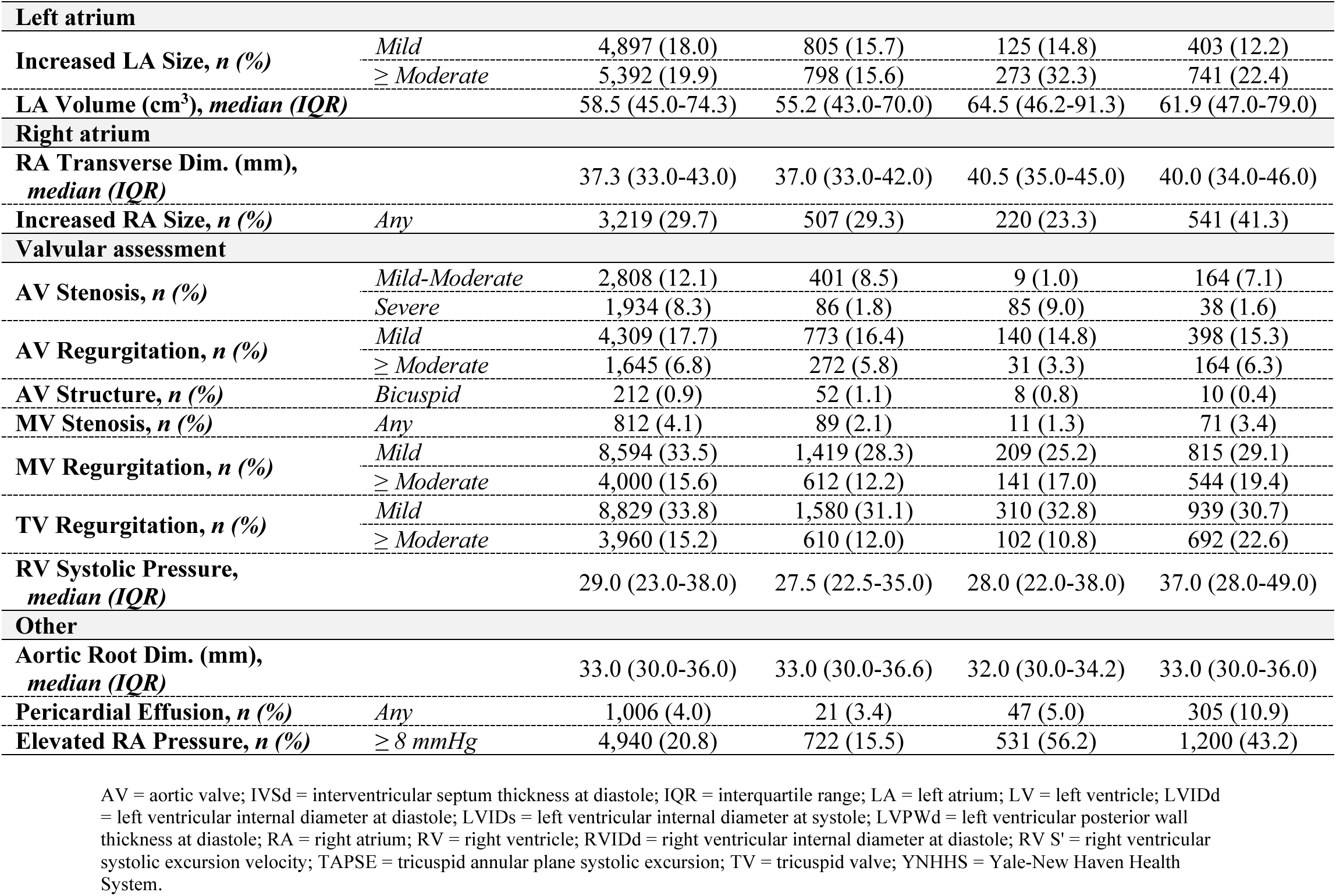
Description of Internal and External Validation Cohorts.

The external RVENet+ cohort included 18,862 echocardiographic videos from 944 complete echocardiographic studies of 831 patients from the Heart and Vascular Center of Semmelweis University in Budapest, Hungary from November 2013-March 2021 (median [IQR] age: 50.0 [25.0-66.0] years, 586 [62.1%] males, 944 [100%] reported White). This cohort featured a unique composition of seven subpopulations, with 57.5% (52.2-61.4) median (IQR) LVEF, 122 (12.9%) studies with moderate or higher LV systolic dysfunction, 85 (9.0%) severe aortic stenosis, 141 (17.0%) moderate or higher mitral regurgitation, etc. Finally, the POCUS cohort consisted of 25,407 videos from 3,310 studies of 3,170 patients undergoing cardiac-focused point-of-care echocardiography in YNHHS-affiliated EDs from September 2015-November 2023 (median [IQR] age: 66.0 [53.0-79.0] years, 1,811 [54.8%] males, 1,122 [35.4%] reported non-White) (**Table 1**). Full description and statistics of all cohorts can be found in **eTable 2**.

### Validation on Diagnostic Echocardiography

PanEcho performed 18 diagnostic classification tasks with 0.91 (IQR: 0.88-0.93) median AUC in internal YNHHS validation and 17 diagnostic tasks with 0.91 (0.85-0.94) median AUC in international RVENet+ validation (**Figure 2**, **eTable 3**). The model accurately assessed ventricular structure and function, with internal AUCs of 0.94 (95% CI: 0.93-0.95) [external: 0.98 (0.98-0.99)] for moderate or higher increased LV size, 0.98 (0.98-0.99) [0.99 (0.98-0.99)] for moderate or higher LV systolic dysfunction, 0.92 (0.91-0.93) [0.92 (0.90-0.93)] for moderate or higher LV diastolic dysfunction, 0.88 (0.86-0.90) [0.98 (0.97-0.99)] for LV wall motion abnormalities, and 0.94 (0.91-0.94) [0.94 (0.92-0.96)] for RV systolic dysfunction. PanEcho also achieved excellent discrimination of valvular disease, with internal AUCs of 0.98 (0.98-0.99) [1.00 (0.99-1.00)] for severe aortic stenosis, 0.96 (0.94-0.98) [1.00 (1.00-1.00)] for mitral stenosis, 0.91 (0.90-0.91) [0.92 (0.91-0.94)] for moderate or higher mitral regurgitation, and 0.90 (0.89-0.91) [0.91 (0.90-0.93)] for moderate or higher tricuspid regurgitation. Additional phenotypes such as pericardial effusion and LV outflow tract obstruction were classified with internal AUCs of 0.91 (0.83-0.97) [0.94 (0.89-0.98)] and 0.94 (0.88-0.89), respectively.

**Figure 2.**
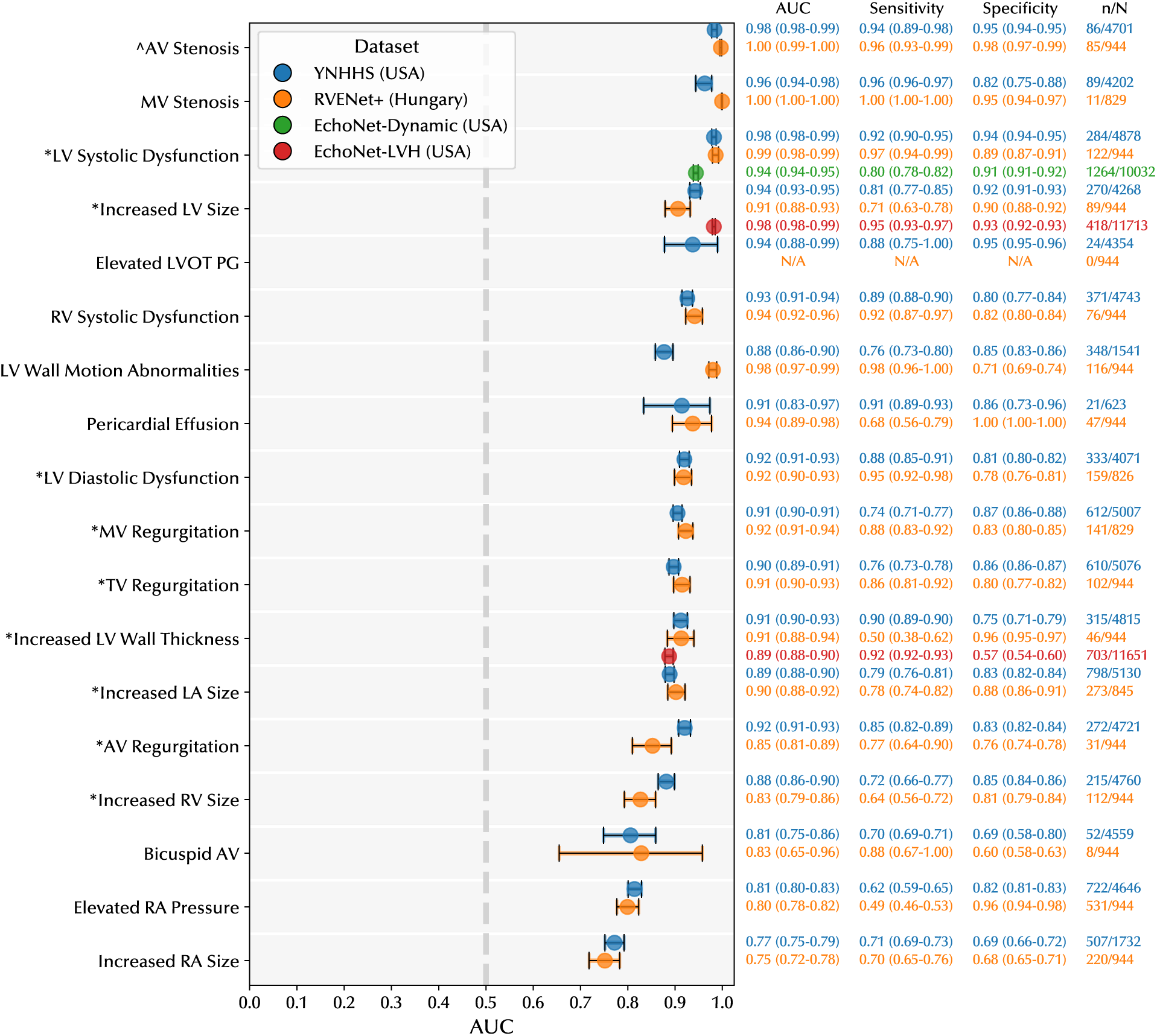
Multi-task Classification Performance Evaluation. Multi-task international validation of PanEcho on diagnostic classification tasks. Results are presented on the internal YNHHS validation cohort (blue) and the external RVENet+ (orange), EchoNet-Dynamic (green), and EchoNet-LVH (red) cohorts. Error bars and values in parentheses represent bootstrapped 95% confidence intervals. n = number of studies meeting the label definition; N = number of non-missing labels; * = moderate or higher; = severe. AUC = area under the receiver operating characteristic curve; AV = aortic valve; LA = left atrium; LV = left ventricle; LVOT = left ventricular outflow tract; PG = pressure gradient; RA = right atrium; RV = right ventricle; TV = tricuspid valve; YNHHS = Yale-New Haven Health System.

Beyond categorical classification, PanEcho estimated 21 echocardiographic parameters with 0.13 median normalized MAE (IQR: 0.10-0.18) in internal validation and 0.16 median normalized MAE (0.11-0.23) in the external RVENet+ cohort (**Figure 3, eTable 4)**. The model accurately quantified LV dimensions and function, with MAEs ranging from 4.2% (95% CI: 4.1-4.3) [external: 4.5% (4.3-4.7)] for LVEF to 1.3 mm (1.3-1.3) [1.3 mm (1.3-1.4)] for LV interventricular septum thickness (IVSd), 1.2 mm (1.1-1.2) [1.1 mm (1.0-1.1)] for LV posterior wall thickness (LVPWd), and 3.8 mm (3.7-3.9) [3.7 mm (3.6-3.7)] for LV internal diameter (LVIDd). For the RV, PanEcho estimated RVIDd with 4.0 mm (3.9-4.1) MAE [3.9 mm (3.7-4.0)], tricuspid plane excursion velocity (TAPSE) with 3.4 mm (3.3-3.4) MAE [3.8 mm (3.6-3.9)], and RV systolic excursion velocity (RV S’) with 1.9 cm/s (1.9-2.0) MAE [2.0 cm/s (1.9-2.1)]. Atrial dimensions such as left atrial (LA) internal diameter (LAIDs) and LA volume were estimated with internal MAEs of 4.0 mm (3.9-4.1) [4.0 mm (3.8-4.3)] and 9.4 cm^3^ (9.2-9.6) [13.4 cm^3^ (12.9-13.9)]. Finally, PanEcho quantified Doppler-derived measurements with MAEs of 0.3 m/s (0.3-0.3) [0.4 m/s (0.3-0.4)] for peak aortic velocity, 5.6 mmHg (5.4-5.7) [6.5 mmHg (6.1-6.9)] for tricuspid peak gradient, and 2.0 (1.9-2.0) [2.2 (2.1-2.3)] for mean E/e’ ratio. See **eFigure 1** for Bland-Altman plots comparing key AI vs. ground truth measurements.

**Figure 3.**
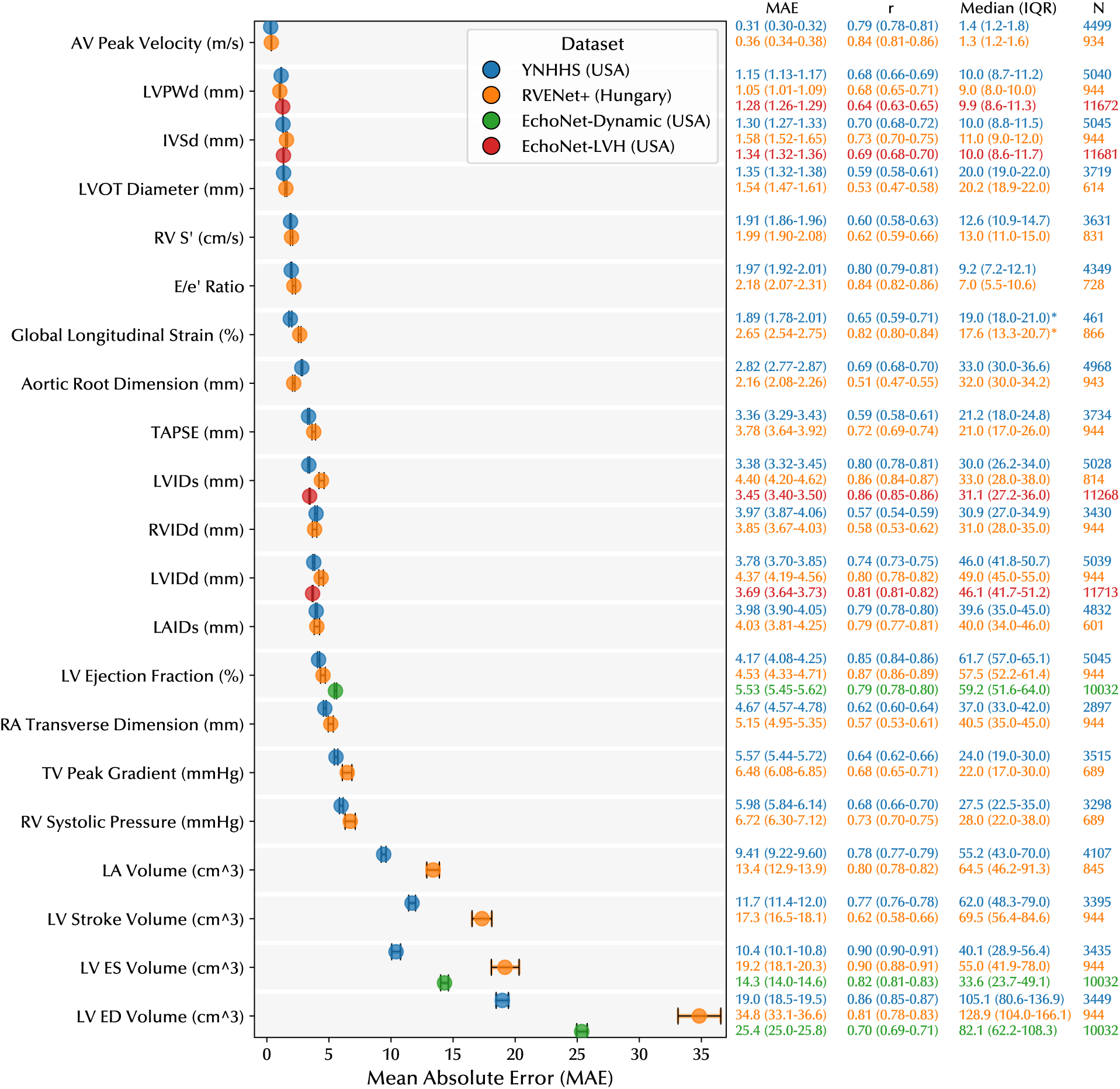
Multi-task Regression Performance Evaluation. Multi-task international validation of PanEcho on continuous parameter estimation tasks. Results are presented on the internal YNHHS validation cohort (blue) and the external RVENet+ (orange), EchoNet-Dynamic (green), and EchoNet-LVH (red) cohorts. Error bars and values in parentheses represent bootstrapped 95% confidence intervals. The “Mean” column represents the mean (SD) value of measurements in each cohort. N = number of non-missing measurements. AV = aortic valve; ED = end-diastolic; ES = end-systolic; IVSd = interventricular septum thickness at diastole; LA = left atrium; LV = left ventricle; LAIDs = left atrial internal diameter at systole; LVIDd = left ventricular internal diameter at diastole; LVIDs = left ventricular internal diameter at systole; LVOT = left ventricular outflow tract; LVPWd = left ventricular posterior wall thickness at diastole; RA = right atrium; RV = right ventricle; RVIDd = right ventricular internal diameter at diastole; RV S’ = right ventricular systolic excursion velocity; TV = tricuspid valve; YNHHS = Yale-New Haven Health System.

In addition to accuracy, PanEcho demonstrated interpretability by correctly identifying the most relevant views for each task (**eFigure 2**), fairness by exhibiting demographic parity across sex and race (**eFigure 3**), and robustness by reliably estimating LVEF under varying image quality (**eFigure 4**). A full description of these auxiliary analyses can be found in the **eMethods**.

### Validation on Point-of-care Echocardiography

PanEcho performed 15 diagnostic classification tasks with 0.91 median AUC (IQR: 0.87-0.94) in an abbreviated TTE cohort, as well as 14 tasks with 0.85 (0.77-0.87) median AUC in a real-world POCUS cohort of bedside ED acquisitions (**Figure 4, eTable 5**). Ventricular assessment remained strong in these limited settings, with AUCs of 0.98 (95% CI: 0.97-0.98) for moderate or higher LV systolic dysfunction in the simulated point-of-care cohort [POCUS: 0.93 (0.92-0.94)], 0.94 (0.92-0.95) [0.89 (0.87-0.90)] for moderate or higher increased LV size, 0.91 (0.89-0.92) [0.83 (0.82-0.85)] for moderate or higher LV diastolic dysfunction, 0.93 (0.91-0.94) [0.85 (0.83-0.87)] for moderate or higher increased LV wall thickness, 0.92 (0.90-0.93) [0.85 (0.84-0.86)] for RV systolic dysfunction, and 0.88 (0.86-0.90) [0.87 (0.86-0.89)] for moderate or higher increased RV size. PanEcho also accurately detected severe aortic stenosis with 0.96 (0.94-0.97) AUC [0.86 (0.81-0.92)] and mitral stenosis with 0.94 (0.92-0.96) [0.92 (0.88-0.95)] from simplified acquisitions. Additional diagnostic findings of pericardial effusion and elevated LV outflow tract pressure were detected with AUCs of 0.91 (0.82-0.97) [0.86 (0.84-0.88)] and 0.94 (0.87-0.99), respectively.

**Figure 4.**
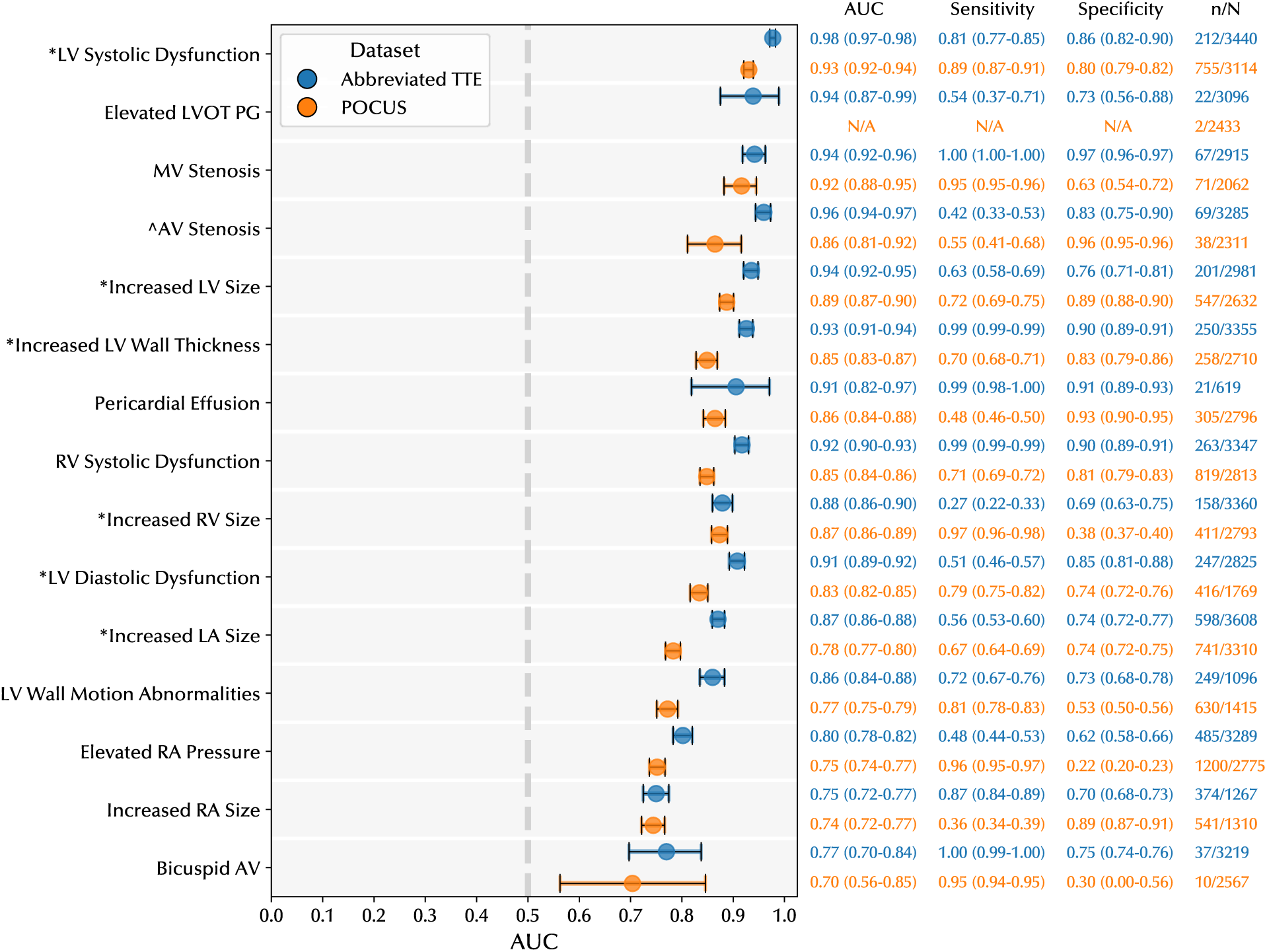
Performance Evaluation at the Point of Care. Multi-task validation of PanEcho on diagnostic classification tasks in a simulated and real-world POCUS dataset. YNHHS Five-View refers to evaluation on the internal YNHHS validation set, except only using up to one video from the following key views per study: PLAX, PSAX, A4C, A5C, A2C. Error bars and values in parentheses represent bootstrapped 95% confidence intervals. Grey dashed line represents the performance of random guessing. n = number of studies meeting the label definition; N = number of non-missing labels; * = moderate or higher, = severe. A2C = apical 2-chamber; A3C = apical 3-chamber; A4C = apical 4-chamber; A5C = apical 5-chamber; AUC = area under the receiver operating characteristic curve; AV = aortic valve; LA = left atrium; LV = left ventricle; LVOT = left ventricular outflow tract; PG = pressure gradient; RA = right atrium; PLAX = parasternal long axis; PSAX = parasternal short axis; RV = right ventricle; TV = tricuspid valve; YNHHS = Yale-New Haven Health System.

## DISCUSSION

We present PanEcho, a view-agnostic deep learning model for automated echocardiography interpretation developed and validated on over one million videos spanning a broad range of acquisitions and phenotypes. PanEcho advances the current state-of-the-art in AI-enabled echocardiography, enabling simultaneous estimation of all key parameters of cardiac structure and function from any combination of 2D views. The model integrates multi-view information from complete echocardiographic studies while maintaining accurate reporting on simplified imaging protocols and technically limited POCUS acquisitions from the ED. PanEcho is open-source, extensively validated internationally, and at the point of care, and may signal a shift from specialized single-task models toward generalized multi-task AI models poised for clinical deployment.

PanEcho was developed to address a critical gap in AI for echocardiography: the predominance of single-view, single-task models. Unlike prior approaches that require a particular echocardiographic view or sequence,^12,14,16,24,31^ PanEcho flexibly integrates videos from all available 2D views. Further, while prior work has primarily been limited to specialized single-task models,^8,13,14,20,22–24^ PanEcho performs end-to-end estimation of all variables forming the core of a complete echocardiographic report. This unified approach is closer to clinical practice – where a cardiologist synthesizes multiple views and imaging modes to assess all aspects of cardiovascular health – and more efficiently scales to clinical deployment. In the previous paradigm, single-task models would pose considerable implementation challenges, especially in computationally constrained environments such as on-device deployment on a handheld device for point-of-care use.

To understand the contributions of this study, PanEcho should be evaluated in the context of state-of-the-art commercial systems^16,31^ and academic efforts.^25,32^ While all report video-based AI models that are capable of automating many aspects of echocardiographic reporting, key differences exist in accessibility and extent of validation. First, unlike commercial products, PanEcho is fully open-source, accelerating research on AI for echocardiography. Second, PanEcho is evaluated across 39 diverse labels spanning key diagnostic findings in a complete echocardiographic report, while comparable studies validate only on subsets of these labels. Finally, PanEcho has been internationally validated across multiple sites on both complete diagnostic-quality studies and technically limited acquisitions by non-expert operators, in contrast to prior work. This extensive validation enables unique opportunities for advancing diagnostic care in point-of-care settings and community clinics, where access to equipment and personnel needed for complete diagnostic echocardiography is limited.

There are three primary clinical applications for PanEcho: (i) as a preliminary reader for assisted interpretation in echocardiography labs, (ii) as a tool to identify potentially missed abnormalities in existing echocardiography databases, and (iii) as an efficient screening system for accelerated protocols and point-of-care exams performed by non-experts. Given its excellent performance on complete echocardiograms, PanEcho can support expert readers by identifying key abnormalities for further interrogation. Prospective integration into a real-world echocardiography lab workflow is needed to evaluate factors such as interface design and the effect of human-AI collaboration on interpretation speed and accuracy. PanEcho may also be used to efficiently parse existing repositories of echocardiograms for potential clinical abnormalities that were missed at the time of examination to be flagged for review. Finally, PanEcho is uniquely suited for point-of-care settings and community clinics given its extensive validation on POCUS acquisitions. Given the scarcity of echocardiographic expertise^33,34^ and increasing accessibility of portable ultrasound devices,^6,7,35^ PanEcho could provide cardiovascular healthcare that might otherwise be inaccessible to many communities. Future work to validate this approach includes prospective human vs. PanEcho evaluation on point-of-care scans from non-sonographers.

### Limitations

Certain limitations merit consideration. First, our model is limited to 2D grayscale and color Doppler echocardiographic videos, excluding other acquisitions such as still frames, spectral Doppler, strain imaging, and 3D echocardiography, which could improve the predictive performance and versatility of PanEcho. Second, unlike other approaches,^13,15–17,31^ our method does not incorporate a segmentation step for echocardiographic measurements. While our direct estimation approach yields strong numerical results, it may limit the model’s interpretability; future iterations of PanEcho may supplement our current approach with a segmentation overlay on select views for improved transparency and ease of use in clinical deployment. Systematic differences in measurement methods^36^ limit performance on tasks such as LV volumes, where RVENet+ labels were derived from 3D echocardiography vs. the mixture of 2D and 3D methods used in routine practice across other cohorts. Similarly, performance on certain labels – such as bicuspid aortic valve and right atrial abnormalities – is limited by low prevalence^37^ and difficulty of interpretation for human experts,^38–40^ causing class imbalance and noisy ground truth labels, respectively. Finally, PanEcho is limited to the specific suite of 39 tasks as defined in this study and, for certain tasks, may fail to provide sufficiently granular interpretations to independently guide patient care. For this reason, a PanEcho-assisted workflow would still ultimately rely on expert supervision to ensure reliable diagnosis and treatment based on the broader clinical context of each patient beyond echocardiography.

## CONCLUSION

We report the development and validation of an AI system that automatically interprets key aspects of echocardiograms – spanning ventricular structure and function to valvular disease and more – maintaining high accuracy across geography and time from complete and technically limited studies. PanEcho and similar tools may be used as an adjunct reader in echocardiography labs or rapid AI-enabled screening tools in point-of-care settings.

## DATA AVAILABILITY

The YNHHS data used in this study is not available for public sharing due to the restrictions in our IRB agreement. However, deidentified validation data may be made available to researchers under a data use agreement upon publication. While the external RVENet+ dataset has not been approved for release, the original RVENet data is available through https://rvenet.github.io/dataset/. The external EchoNet-Dynamic and EchoNet-LVH datasets can be accessed through the Stanford AIMI Shared Datasets repository at the following links, respectively: https://stanfordaimi.azurewebsites.net/datasets/834e1cd1-92f7-4268-9daa-d359198b310a and https://stanfordaimi.azurewebsites.net/datasets/5b7fcc28-579c-4285-8b72-e4238eac7bd1.

## CODE AVAILABILITY

The code repository for this study is available at https://github.com/CarDS-Yale/PanEcho.

## ACKNOWLEDGMENTS

We thank Andreas Coppi, PhD (Section of Cardiovascular Medicine, Department of Internal Medicine, Yale School of Medicine, New Haven, CT, USA) for instrumental work facilitating the external validation of PanEcho in the RVENet+ cohort. We also gratefully acknowledge Béla Merkely, MD, PhD (Heart and Vascular Center, Semmelweis University, Budapest, Hungary) for his leadership in the establishment and sustained development of the RVENet+ dataset.

## REFERENCES

1. Mitchell C, Rahko PS, Blauwet LA, et al. Guidelines for performing a comprehensive transthoracic echocardiographic examination in adults: Recommendations from the American society of echocardiography. J Am Soc Echocardiogr. 2019;32(1):1–64.

2. Wei C, Milligan M, Lam M, Heidenreich PA, Sandhu A. Variation in cost of echocardiography within and across United States hospitals. J Am Soc Echocardiogr. 2023;36(6):569–577.e4.

3. Virnig BA, Shippee ND, O’Donnell B, Zeglin J, Parashuram S. Trends in the Use of Echocardiography, 2007 to 2011. Agency for Healthcare Research and Quality (US); 2014.

4. Pillai B, Salerno M, Schnittger I, Cheng S, Ouyang D. Precision of echocardiographic measurements. J Am Soc Echocardiogr. 2024;37(5):562–563.

5. He B, Kwan AC, Cho JH, et al. Blinded, randomized trial of sonographer versus AI cardiac function assessment. Nature. 2023;616(7957):520–524.

6. Díaz-Gómez José L., Mayo Paul H., Koenig Seth J. Point-of-Care Ultrasonography. N Engl J Med. 2021;385(17):1593–1602.

7. Narang A, Bae R, Hong H, et al. Utility of a Deep-Learning Algorithm to Guide Novices to Acquire Echocardiograms for Limited Diagnostic Use. JAMA Cardiol. 2021;6(6):624–632.

8. Holste G, Oikonomou EK, Mortazavi BJ, et al. Severe aortic stenosis detection by deep learning applied to echocardiography. Eur Heart J. Published online August 23, 2023. doi:10.1093/eurheartj/ehad456

9. Krishna H, Desai K, Slostad B, et al. Fully Automated Artificial Intelligence Assessment of Aortic Stenosis by Echocardiography. J Am Soc Echocardiogr. 2023;36(7):769–777.

10. Huang Z, Long G, Wessler B, Hughes MC. A New Semi-supervised Learning Benchmark for Classifying View and Diagnosing Aortic Stenosis from Echocardiograms. In: Jung K, Yeung S, Sendak M, Sjoding M, Ranganath R, eds. Proceedings of the 6th Machine Learning for Healthcare Conference. Vol 149. Proceedings of Machine Learning Research. PMLR; 06--07 Aug 2021:614–647.

11. Oikonomou EK, Holste G, Yuan N, et al. A Multimodality Video-Based AI Biomarker For Aortic Stenosis Development And Progression. JAMA Card. 2024;9(6):534–544.

12. Zhang J, Gajjala S, Agrawal P, et al. Fully Automated Echocardiogram Interpretation in Clinical Practice. Circulation. 2018;138(16):1623–1635.

13. Ouyang D, He B, Ghorbani A, et al. Video-based AI for beat-to-beat assessment of cardiac function. Nature. 2020;580(7802):252–256.

14. Ghorbani A, Ouyang D, Abid A, et al. Deep learning interpretation of echocardiograms. NPJ Digit Med. 2020;3:10.

15. Reddy CD, Lopez L, Ouyang D, Zou JY, He B. Video-Based Deep Learning for Automated Assessment of Left Ventricular Ejection Fraction in Pediatric Patients. J Am Soc Echocardiogr. 2023;36(5):482–489.

16. Tromp J, Bauer D, Claggett BL, et al. A formal validation of a deep learning-based automated workflow for the interpretation of the echocardiogram. Nat Commun. 2022;13(1):6776.

17. Zeng Y, Tsui PH, Pang K, et al. MAEF-Net: Multi-attention efficient feature fusion network for left ventricular segmentation and quantitative analysis in two-dimensional echocardiography. Ultrasonics. 2023;127:106855.

18. Khera R, Oikonomou EK, Nadkarni GN, et al. Transforming Cardiovascular Care With Artificial Intelligence: From Discovery to Practice: JACC State-of-the-Art Review. J Am Coll Cardiol. 2024;84(1):97–114.

19. Goto S, Mahara K, Beussink-Nelson L, et al. Artificial intelligence-enabled fully automated detection of cardiac amyloidosis using electrocardiograms and echocardiograms. Nat Commun. 2021;12(1):2726.

20. Long A, Haggerty CM, Finer J, et al. Deep Learning for Echo Analysis, Tracking, and Evaluation of Mitral Regurgitation (DELINEATE-MR). Circulation. Published online June 17, 2024. doi:10.1161/CIRCULATIONAHA.124.068996

21. Ferreira DL, Salaymang Z, Arnaout R. Label-free segmentation from cardiac ultrasound using self-supervised learning. arXiv [eessIV]. Published online October 10, 2022. http://arxiv.org/abs/2210.04979

22. Vrudhula A, Duffy G, Vukadinovic M, Liang D, Cheng S. High Throughput Deep Learning Detection of Mitral Regurgitation. medRxiv. Published online 2024. https://www.medrxiv.org/content/10.1101/2024.02.08.24302547.abstract

23. Vrudhula A, Vukadinovic M, Haeffle C, et al. Deep Learning Phenotyping of Tricuspid Regurgitation for Automated High Throughput Assessment of Transthoracic Echocardiography. medRxiv. Published online June 24, 2024. doi:10.1101/2024.06.22.24309332

24. Duffy G, Cheng PP, Yuan N, et al. High-Throughput Precision Phenotyping of Left Ventricular Hypertrophy With Cardiovascular Deep Learning. JAMA Cardiol. 2022;7(4):386–395.

25. Christensen M, Vukadinovic M, Yuan N, Ouyang D. Vision–language foundation model for echocardiogram interpretation. Nat Med. 2024;30(5):1481–1488.

26. Holste G, Oikonomou EK, Mortazavi BJ, Wang Z, Khera R. Efficient deep learning-based automated diagnosis from echocardiography with contrastive self-supervised learning. Commun Med. 2024;4(1):133.

27. Magyar B, Tokodi M, Soós A, et al. RVENet: A large echocardiographic dataset for the deep learning-based assessment of right ventricular function. In: Lecture Notes in Computer Science. Lecture notes in computer science. Springer Nature Switzerland; 2023:569–583.

28. Tokodi M, Magyar B, Soós A, et al. Deep Learning-Based Prediction of Right Ventricular Ejection Fraction Using 2D Echocardiograms. JACC Cardiovasc Imaging. 2023;16(8):1005–1018.

29. Oikonomou EK, Vaid A, Holste G, et al. Artificial intelligence-guided detection of under-recognised cardiomyopathies on point-of-care cardiac ultrasonography: a multicentre study. Lancet Digit Health. 2025;7(2):e113–e123.

30. Vaswani A, Shazeer NM, Parmar N, et al. Attention is All you Need. Adv Neural Inf Process Syst. Published online June 12, 2017:5998–6008.

31. Tromp J, Seekings PJ, Hung CL, et al. Automated interpretation of systolic and diastolic function on the echocardiogram: a multicohort study. Lancet Digit Health. 2022;4(1):e46–e54.

32. Vukadinovic M, Tang X, Yuan N, et al. EchoPrime: A multi-video view-informed vision-language model for comprehensive echocardiography interpretation. arXiv [csCV]. Published online October 12, 2024. http://arxiv.org/abs/2410.09704

33. Narang A, Sinha SS, Rajagopalan B, et al. The supply and demand of the cardiovascular workforce: Striking the right balance. J Am Coll Cardiol. 2016;68(15):1680–1689.

34. Colebourn CL. Future-proofing UK echocardiography. Br J Cardiol. 2023;30(4):36.

35. Ginsburg AS, Liddy Z, Khazaneh PT, May S, Pervaiz F. A survey of barriers and facilitators to ultrasound use in low- and middle-income countries. Sci Rep. 2023;13(1):1–11.

36. Dorosz JL, Lezotte DC, Weitzenkamp DA, Allen LA, Salcedo EE. Performance of 3-dimensional echocardiography in measuring left ventricular volumes and ejection fraction: a systematic review and meta-analysis. J Am Coll Cardiol. 2012;59(20):1799–1808.

37. Sillesen A, Vøgg O, Pihl C, et al. Prevalence of bicuspid aortic valve and associated aortopathy in newborns in Copenhagen, Denmark. JAMA. 2021;325(6):561–567.

38. Jain R, Ammar KA, Kalvin L, et al. Diagnostic accuracy of bicuspid aortic valve by echocardiography. Echocardiography. 2018;35(12):1932–1938.

39. Sun ZY, Li Q, Li J, Zhang MW, Zhu L, Geng J. Echocardiographic evaluation of the right atrial size and function: Relevance for clinical practice. Am Heart J Plus. 2023;27(100274):100274.

40. Grünig E, Henn P, D’Andrea A, et al. Reference values for and determinants of right atrial area in healthy adults by 2-dimensional echocardiography. Circ Cardiovasc Imaging. 2013;6(1):117–124.

