## Supplement for "PanEcho: Complete AI-enabled echocardiography interpretation with multi-task deep learning"

### eMethods

#### Echocardiography Data Extraction

As per our previously published echocardiography processing pipeline,[1] pixel data from 2D echocardiographic videos was extracted from the raw Digital Imaging and Communications in Medicine (DICOM) files and deidentified by masking out peripheral pixels containing protected health information. All videos were processed by a pretrained view classifier[2] to determine both the echocardiographic view and imaging mode by randomly selecting ten frames and averaging predicted view probabilities over the ten frames. While the view classifier could discriminate 23 fine-grained view variations, we considered the following key views for later view relevance analysis: apical 2-, 3-, 4-, and 5-chamber (A2C, A3C, etc.), parasternal long axis (PLAX), parasternal short axis (PSAX), right ventricle (RV) inflow, subcostal, and suprasternal. These steps were applied to all YNHHS, RVENet+, and POCUS cohorts.

#### Automated Color Doppler Detection

To detect the use of color Doppler imaging in the YNHHS cohort, we performed a three-step process of identifying videos that were (i) classified as "Other" by the view classifier, (ii) classified as color Doppler by a custom color Doppler detection model, and (iii) contained a nontrivial amount of red pixels.

For step (ii), we developed a dedicated color Doppler detection model on a manually curated dataset of echocardiogram frames derived from studies not present in the YNHHS dataset used for PanEcho development. Specifically, we manually labeled the presence of color Doppler in all videos from five studies and included videos from another five studies that were known to not contain color Doppler as determined by the view classifier. This dataset of 11,240 labeled frames was then randomly split intro training (80%) and validation (20%) sets at the study level in order to ensure sufficient sample size and diversity of labeled examples in the held-out validation set. An ImageNet-pretrained ConvNeXt-T[3] convolutional neural network (CNN) was trained to classify the presence of color Doppler using a batch size of 128, the Adam optimizer[4] with a learning rate of 0.0001, and a weighted binary cross-entropy loss for ten epochs. All frames were downsampled to 256 x 256 resolution, center cropped to 224 x 224, and normalized with ImageNet channel-wise means and standard deviations. The model achieved 100% accuracy on the validation set and was then applied to all videos classified as "Other" by the view classifier. Similar to view classification, ten randomly selected frames from each video were passed to the color Doppler detection model, and predictions were averaged over the ten frames; videos classified as "Other" but not classified as color Doppler at this step were excluded from the cohort.

As a final quality check, for step (iii), the candidate color Doppler videos underwent color detection to assert the presence of the hue of red typically present in color Doppler echocardiography to indicate blood flow toward the transducer. Frames in each video were converted to the HSV color space, and individual pixels were determined to be red if their HSV values fell between (-10, 150, 150) and (10, 255, 255). Videos were deemed to contain a nontrivial number of red pixels if the total fraction of unique pixels that were red at any point in the video exceeded 1%; videos that passed the previous two steps but failed this step were discarded. Beyond filtering out videos that were neither color Doppler nor 2D B-mode, we did not perform any further quality control to encourage robustness to variations in acquisition quality (e.g., low-contrast or off-axis images), ultrasound machine settings, etc. encountered in real-world clinical practice.

To detect color Doppler in the RVENet+ and POCUS cohorts, we only applied steps (i) and (iii) outlined above for simplicity; the intermediate step (ii) above was needed to obtain reliable color Doppler labels for later view relevance analysis, which was only performed in the YNHHS cohort. For the POCUS cohort, all videos with color Doppler imaging were removed, as this is not a standard acquisition in a cardiac-focused point-of-care ultrasound examination.

#### Cohort Curation

After color Doppler detection, we limited the YNHHS cohort to contain at most four unique studies per patient – randomly selecting four studies to keep for patients examined at least five times – to prevent overrepresentation of specific patients and outcomes. Next, the resulting dataset was split into development and internal validation sets, with studies performed from July to December 2022 set aside as a temporally distinct test set. The remaining studies from January 2016 to June 2022 were to be used for model development after removing studies from all patients present in the test set to prevent data leakage. The development set was randomly partitioned into training (92.5%) and validation (7.5%) sets at the patient level for model training.

Since the POCUS cohort consisted of emergency department acquisitions by non-experts, greater quality control was needed to ensure minimal diagnostic quality. First, non-cardiac videos from linear array transducers were detected and removed by identifying videos with significant non-background content in the upper corners. Second, in addition to removing color Doppler acquisitions as described above, all subcostal and suprasternal views were removed as these often did not capture the heart. Finally, only studies with at least three of the following five views represented in the study were retained: PLAX, PSAX at the level of the papillary muscle, A4C, A2C, and A5C. This was done to ensure the inclusion of studies following a basic limited acquisition protocol with key views necessary for the multi-task analysis performed by PanEcho.

**Echocardiography Data Preprocessing**

In all cohorts, all videos underwent thorough deidentification by masking out pixels beyond the central image content. Namely, each frame was binarized with a fixed threshold, then all pixels outside the convex hull of the largest contour were masked out. All videos were then cropped to the central image content in a temporally consistent manner and downsampled to 256 x 256 resolution with bicubic interpolation. These steps were not necessary for the EchoNet-Dynamic and EchoNet-LVH datasets, which were already deidentified; EchoNet-LVH videos were downsampled to 256 x 256 resolution with bicubic interpolation, and EchoNet-Dynamic videos – already downsampled to 112 x 112 resolution – were upsampled to 256 x 256 resolution with bilinear interpolation to maintain consistent input spatial resolution.

**Echocardiographic Reporting Labels**

The labels used in this study were determined by reviewing all standard structured echocardiography fields included in the data reporting software (Lumedx®, Oakland, CA) used by YNHHS hospitals. Our clinical team compared these fields against elements from a standard ASE report and identified clinically relevant labels that were consistently captured in YNHHS practice and would provide comprehensive coverage of myocardial size, function, thickness spanning all chambers, as well as valve function and structure and miscellaneous features commonly reported in TTE examinations (e.g., pericardial effusion, great vessels, etc.). We excluded labels where missingness was so extreme that sample sizes would not be sufficient for reliable evaluation (<30 total positive examples for classification tasks) or where missingness was decidedly not-at-random. For instance, we excluded aortic regurgitation pressure half-time, which was generally only reported when considered relevant to patient care.

For YNHHS and POCUS studies, labels were extracted from the local electronic echocardiography reporting system (Lumedx®, Oakland, CA). For the RVENet+ dataset, labels were extracted from the local picture archiving and communication system (Philips Ultrasound Workspace, Philips Medical Systems, Best, the Netherlands) of the Echocardiography Research Laboratory at the Heart and Vascular Center of Semmelweis University; LV volumes and EF were measured using 3D echocardiography (LV-Analysis 3, TomTec Imaging Systems, Unterschleissheim, Germany). For all cohorts, labels reflect the final measurements and reporting confirmed by certified echocardiographers. See **eTable 1** for a description of the guidelines used to determine each categorical diagnostic label; while these guidelines reflect standard practice, the final reporting is ultimately subject to the discretion of the original interpreting cardiologist given the broader clinical context of the patient.

To minimize the effect of extreme outliers on regression tasks, we applied winsorization to all continuous variables, limiting the lowest and highest values to the 0.5 and 99.5 percentile values, respectively. These cutoffs were determined based on the ground truth measurements present in the YNHHS training set and applied to all other cohorts accordingly. Additionally, given the relatively low prevalence of severe phenotypes across certain categorical labels in classification tasks, we pooled moderate and severe phenotypes into shared severity groups for selected tasks. This was done to ensure sufficient sample sizes for reliable model development and evaluation on these rare classes. In the YNHHS and POCUS validation datasets, erroneous LV ejection fraction values were removed (deemed missing) by identifying values entered as 27.5% but having LV Systolic Dysfunction entered as "Mild" or "None".

For the EchoNet-Dynamic and EchoNet-LVH cohorts, a total of 10 labels were extracted based on the measurements provided in these publicly available datasets. EchoNet-Dynamic provided LV ejection fraction, LV ED Volume, and LV ES Volume measurements for each video (one provided per study). Based on this, we determined categorical LV Systolic Dysfunction labels as follows: "None" = LVEF ≥ 54%, "≥ Moderate" = LVEF ≤ 40%, and "Mild" otherwise. EchoNet-LVH provided LVIDd, LVIDs, LVPWd, and IVSd measurements. From these, we determined categorical Increased LV Size labels via ""≥ Moderate" = LVIDd ≥ 6.4 cm, "Normal" = LVIDd ≤ 5.2 cm, and "Mild" otherwise, as well as Increased LV Wall Thickness via ""≥ Moderate" = IVSd ≥ 1.3 cm & LVPWd ≥ 1.3 cm, "Any" = IVSd ≥ 1.1 cm & LVPWd ≥ 1.1 cm, and "None" otherwise. While standard cutoffs for these conditions are sex-dependent, these conservative thresholds were chosen since patient sex was not provided in EchoNet-LVH nor EchoNet-Dynamic.

**PanEcho Implementation Details**
In designing the architecture for PanEcho, we adopted a decoupled "2+1D" approach to modeling echocardiogram videos – with separate modules to learn spatial and temporal features – primarily for downstream flexibility; for instance, our 2D image backbone can be readily adapted for any echocardiographic task, while a 3D backbone would be more difficult to retrofit to a 2D image-only task such as segmentation. Each video frame is first processed by the 2D image encoder, an ImageNet[5]-pretrained ConvNeXt-T[3] convolutional neural network (CNN), which produces a learned feature vector, or representation, of each frame. These frame-wise representations are then interpreted as an ordered sequence – like words in a sentence in natural language processing – and modeled using self-attention[6] to learn time-varying associations over the frames. Frame order is embedded via sinusoidal positional encoding, which is then elementwise added to the frame-wise feature vectors and fed to a Transformer encoder consisting of four layers, each with eight self-attention heads. Mean pooling is then used to aggregate frame-wise feature vectors into a single video-level representation, which is used as input to the task-specific output heads. Each output head consists of a Dropout[7] layer with probability 0.25 and a fully-connected layer. Both regression and binary classification tasks used one output neuron, the latter followed by a sigmoid activation. Multi-class classification tasks with $k$ classes used $k$ output neurons with softmax activation, and multi-label classification tasks were modeled with separate binary classification heads for each class. PanEcho was trained to minimize the mean of all valid task-specific losses – cross-entropy for classification tasks and mean squared error for regression tasks. Studies with missing labels were masked from task-specific loss computation, and tasks with fully missing labels (i.e., no studies in the mini-batch had ground truth labels) were masked from aggregated loss computation. To control for varying units and scales of regression tasks, we first divided each regression loss by the mean observed value of that measurement in the training set before loss aggregation.

PanEcho was implemented and trained in PyTorch[8] version 2.1.0 with distributed training across eight NVIDIA A100 graphics processing units (GPUs) with automatic mixed precision to maximize throughput. During training, the model received as input a randomly sampled video clip of 16 consecutive frames from an echocardiogram, following prior work.[1] To increase robustness to variations in acquisition and increase effective sample size, the following augmentations were performed to all video frames in a temporally consistent manner: random crop to 224 x 224 resolution, random horizontal flip with probability 0.5, random rotation within (-15°, 15°), then followed by ImageNet normalization. The model was trained with a batch size of 16 per GPU, the Adam optimizer[4], and minimized the multi-task loss described above with learning rate 0.0001. The learning rate was reduced by a factor of 0.5 if the validation metric (mean classification AUC and regression $R^2$ across all tasks) did not improve for three consecutive epochs; though MAE was the primary evaluation metric for regression tasks, this validation metric was chosen because AUC and $R^2$ are both increasing and bounded to $[0, 1]$. The model was trained for a maximum of 30 epochs with early stopping if validation metric did not improve for 10 consecutive epochs. At test time, four 16-frame clips are randomly sampled from each video and task-wise predictions are averaged over all clips to produce video-level predictions. Since PanEcho is view-agnostic and labels are determined at the study level, predictions from all videos acquired during the same study (regardless of imaging mode or view) were averaged to form a single study-level prediction for each task.

**Validation on Diagnostic Echocardiography**
PanEcho was evaluated on all available labels in the internal YNHHS validation set (39) and the external RVENet+ (38), EchoNet-Dynamic (4), and EchoNet-LVH (6) cohorts. While all 39 labels were extracted for the RVENet+ cohort, there were no instances of Elevated LVOT PG in the cohort, meaning performance metrics could not be computed for this task. As the YNHHS and RVENet+ were multi-view datasets including all available 2D echocardiographic views, video-level predictions for all acquisitions in a study were averaged to produce holistic study-level predictions for each task at test time. EchoNet-Dynamic and EchoNet-LVH only provided individual videos – from the A4C and PLAX views, respectively – meaning PanEcho performance was determined based on one video per study.

**Validation on Point-of-care Echocardiography**
PanEcho was additionally evaluated on a simulated abbreviated TTE dataset and a real-world POCUS dataset from bedside acquisitions by non-experts. The Abbreviated TTE dataset was derived from the internal YNHHS validation set by removing all color Doppler videos and selecting at most one videos from the following five key views per study: PLAX, PSAX at the level of the papillary muscle, A4C, A5C, and A2C. While the view classifier was capable of discriminating variations of several view types (e.g., A4C vs. A4C – LV occluded vs. A4C – LA occluded), only videos from the "primary" view were considered for this analysis. In the case that multiple videos

from one of the five key views were present in a study, the video with the highest predicted view probability was selected.

**Task-specific View Relevance Analysis**
We defined the normalized view relevance score

$$R_{v,t} = \begin{cases} \frac{m_{v,t}}{\max_v (m_{v,t})}, & t \text{ is a classification task} \\ \frac{m_{v,t}}{\min_v (m_{v,t})}, & t \text{ is a regression task} \end{cases},$$

where $R_{v,t}$ is the relevance of view $v$ for task $t$, and $m_{v,t}$ is the performance metric on task $t$ when only using view $v$ (AUC for classification tasks and MAE for regression tasks). This produces a task-normalized score where, for a given task, 1 represents the most informative view, and each score can be interpreted as the "fractional importance relative to the best view." This analysis was performed on the internal YNHHS validation set and metrics were computed after selecting a maximum of three videos per view in a given study with the most confident predicted view probability by the view classifier; this was done to control for the variable prevalence of views – without this, the most common views would be overrepresented within each study and unfairly benefit from a greater ensembling effect after video-level aggregation. For tasks typically performed with or aided by some form of Doppler imaging, we performed this analysis again after including color Doppler videos (from any echocardiographic view) as an additional "view" to assess the task-dependent value of color Doppler imaging.

**eFigure 2** summarizes view relevance for aspects of cardiovascular health by averaging view relevance scores across tasks falling under each category. Specifically, we include the following categories (constituent tasks in parentheses): *Left Ventricular Structure* (IVSd, LVPWd, LVIDs, LVIDd, Increased LV Size [≥ Moderate], Increased LV Wall Thickness [≥ Moderate], LV ED Volume, LV ES Volume), *Left Ventricular Function* (LV Ejection Fraction, LV Stroke Volume, LV Systolic Dysfunction [≥ Moderate], LV Diastolic Dysfunction [≥ Moderate], LV Wall Motion Abnormalities), *Right Ventricle* (RVIDd, TAPSE, RV Systolic Pressure, Increased RV Size [≥ Moderate], RV Systolic Dysfunction), *Left Atrium* (LAIDs, LA Volume, Increased LA Size), *Aortic Valve* (Aortic Root Dimension, AV Structure, AV Stenosis [Severe], AV Regurgitation [≥ Moderate]), *Valvular Regurgitation* (AV Regurgitation [≥ Moderate], MV Regurgitation [≥ Moderate], TV Regurgitation [≥ Moderate]).

**Group Fairness Analysis**
Beyond predictive performance, we also evaluate the demographic parity of PanEcho across protected subgroups of sex and race to assess bias (**eFigure 3**). This analysis involved computation of the difference in AUC (ΔAUC) between subgroups for each classification task in the internal YNHHS validation set. Groupwise comparisons were excluded if the subgroup has <30 positive examples of a particular label to ensure sufficient sample size for reliable evaluation. For comparisons across sex, this meant that analysis could be performed for 15/18 labels; for comparisons across race, analysis could only be performed for 12/18 labels for the White or Caucasian, Black or African American, and Other racial groups. If the 95% CI for ΔAUC overlapped with zero, then PanEcho was considered to demonstrate unbiased predictive performance between those particular subgroups on a given task.

**Image Quality Analysis**
To evaluate PanEcho's robustness to variations in image quality, we assessed its LV ejection fraction estimation performance across images of different quality levels in the internal YNHHS validation set (**eFigure 4**). While there is no standard metric for image quality, we consider the view classifier's predicted view probability – interpreted as its "confidence" – as a proxy for image quality. For this analysis, only videos from the A4C view were included, as this is the key view for LV ejection fraction evaluation. Similar to the view relevance analysis, a maximum of three A4C videos per study were included (selected by maximum view classifier confidence) per study to control for the number of videos used to make study-level predictions. Based on the mean A4C view classifier probability of the ≤3 videos included in each study, five equal-sized bins (quintiles) of image quality were identified: Q1 (0.162, 0.667], Q2 (0.667, 0.847], Q3 (0.847, 0.965], Q4 (0.965, 0.998], and Q5 (0.998, 1]. LV ejection fraction estimation was then evaluated for studies falling within each quintile by MAE.

#### eTable 1 | Diagnostic Label Definitions and Guidelines

| | Class | Suggested Definition |
|---|---|---|
| **AV Regurgitation** | *None, Mild, ≥ Moderate* | Per recommendations[11] |
| **AV Stenosis** | *None, Mild-Moderate, Severe* | Per recommendations[12,13] |
| **AV Structure** | *Bicuspid* | - |
| **Increased LV Size** | *None* | Female: LV EDVi ≤61 $mL/m^2$, LVIDd ≤5.2 cm<br>Male: LV EDVi ≤74 $mL/m^2$, LVIDd ≤5.8 cm |
| | *Mild* | Female: LV EDVi 62-70 $mL/m^2$, LVIDd 5.3-5.6 cm<br>Male: LV EDVi 75-89 $mL/m^2$, LVIDd 5.9-6.3 cm |
| | *≥ Moderate* | Female: LV EDVi ≥71 $mL/m^2$, LVIDd ≥5.7 cm<br>Male: LV EDVi ≥90 $mL/m^2$, LVIDd ≥6.4 cm |
| **Increased LV Wall Thickness** | *Any* | Female: IVSd & LVPWd ≥1.0 cm, alternatively LVMi >95 $g/m^2$<br>Male: IVSd & LVPWd ≥1.1 cm, or LVMi >115 $g/m^2$ |
| | *≥ Moderate* | Female: IVSd & LVPWd ≥1.2 cm; Male: IVSd & LVPWd ≥1.3 cm |
| **LV Systolic Dysfunction** | *None* | Female: LV EF ≥54%; Male: LV EF ≥ 52% |
| | *Mild* | Female: LV EF 41-53%; Male: LV EF 41-51% |
| | *≥ Moderate* | Female & Male: LV EF ≤40% |
| **LV Diastolic Dysfunction** | *None* | Per recommendations[14] |
| | *Mild (~ grade 1)* | |
| | *≥ Moderate (~ grade 2, 3)* | |
| **LV Wall Motion Abnormalities** | *Any* | - |
| **Increased LA Size** | *None* | LAVi≤34 $mL/m^2$ |
| | *Mild* | LAVi 35-41 $mL/m^2$ |
| | *≥ Moderate* | LAVi ≥42 $mL/m^2$ |
| **Elevated LVOT PG** | *≥ 20 mmHg* | - |
| **MV Stenosis** | *Any (≥ Mild)* | Per guidelines[13] |
| **MV Regurgitation** | *None, Mild, ≥ Moderate* | Per guidelines[11] |
| **Pericardial Effusion** | *Any (≥ Mild)* | Excluding trivial effusions, this includes an effusion which is <10 mm but can be seen throughout diastole and systole (moderate 10-20 mm, and large >20 mm) |
| **Elevated RA Pressure** | *≥ 8 mmHg* | E.g., dilated inferior vena cava >2.1 cm with preserved collapsibility >50% with sniff |
| **Increased RA Size** | *Any* | Minor axis length ≥4.6 cm |
| **Increased RV Size** | *None* | RV diameter at mid-ventricle ≤3.5 cm |
| | *Mild* | RV diameter at mid-ventricle 3.6-3.8 cm |
| | *≥ Moderate* | RV diameter at mid-ventricle ≥3.9 cm. For RV EDVi consider ≥160 $mL/m^2$ or RV ESVi ≥ 80 $mL/m^2$ to define moderate or severe dilation. |
| **RV Systolic Dysfunction** | *Any* | RV EF ≤ 40% |
| **TV Regurgitation** | *None, Mild, ≥ Moderate* | Per recommendations[11] |

AV = aortic valve; EDVi = end-diastolic volume index; EF = ejection fraction; ESVi = end-systolic volume index; IVSd = interventricular septal thickness in diastole; LA = left atrium; LAVi = left atrial volume index; LV = left ventricle; LVIDd = left ventricular internal diameter at diastole; LVIDs = left ventricular internal diameter at systole; LVMi = left ventricular mass index; LVOT = left ventricular outflow tract; LVPWd = left ventricular posterior wall thickness in diastole; MV = mitral valve; PG = pressure gradient; RA = right atrium; RV = right ventricle; TV = tricuspid regurgitation.

**eTable 2 | Detailed Description of Study Cohorts**

| | | YNHHS | | | | RVENet+ | POCUS |
|---|---|---|---|---|---|---|---|
| | | **Training** | **Tuning** | **Validation** | **Abbreviated TTE** | | |
| **Dates Sampled** | | 1/2016-6/2022 | 1/2016-6/2022 | 7/2022-12/2022 | 7/2022-12/2022 | 11/2013-3/2021 | 9/2015-11/2023 |
| **Patients** | ***Total**, n* | 18,343 | 1,474 | 4,588 | 3,107 | 831 | 3,170 |
| **Studies** | ***Total**, n* | 25,130 | 2,005 | 5,130 | 3,608 | 944 | 3,310 |
| **Videos** | ***Total**, n* | 999,727 | 79,869 | 114,280 | 9,091 | 18,862 | 25,407 |
| | ***Per Study**, median (IQR)* | 39.0 (30.0-48.0) | 39.0 (31.0-48.0) | 17.0 (12.0-31.0) | 2.0 (1.0-4.0) | 19.5 (17.0-22.0) | 6.0 (5.0-9.0) |
| **Age (years)** | ***Present**, median (IQR)* | 69.0 (58.0-79.0) | 68.0 (58.0-79.0) | 68.0 (57.0-78.0) | 68.0 (57.0-78.0) | 50.0 (25.0-66.0) | 66.0 (53.0-79.0) |
| | ***Missing**, n* | 0 | 0 | 0 | 0 | 0 | 0 |
| **Sex** | ***Male**, n (%)* | 13,184 (52.5) | 1,050 (52.4) | 2,585 (50.4) | 1,846 (51.2) | 586 (62.1) | 1,817 (54.9) |
| | ***Female**, n (%)* | 11,946 (47.5) | 955 (47.6) | 2,545 (49.6) | 1,762 (48.8) | 358 (37.9) | 1,493 (45.1) |
| | ***Missing**, n* | 0 | 0 | 0 | 0 | 0 | 0 |
| **Race** | ***White**, n (%)* | 19,333 (80.8) | 1,541 (81.4) | 3,893 (80.6) | 2,728 (80.0) | 944 (100) | 2,073 (65.8) |
| | ***Black**, n (%)* | 3,455 (14.4) | 256 (13.5) | 679 (14.0) | 501 (14.7) | 0 (0) | 871 (27.7) |
| | ***Asian**, n (%)* | 403 (1.7) | 33 (1.7) | 112 (2.3) | 79 (2.3) | 0 (0) | 68 (2.2) |
| | ***American Indian**, n (%)* | 60 (0.3) | 8 (0.4) | 15 (0.3) | 11 (0.3) | 0 (0) | 12 (0.4) |
| | ***Pacific Islander**, n (%)* | 639 (2.7) | 54 (2.9) | 124 (2.6) | 89 (2.6) | 0 (0) | 115 (3.7) |
| | ***Other**, n (%)* | 36 (0.2) | 0 (0) | 10 (0.2) | 3 (0.1) | 0 (0) | 10 (0.3) |
| | ***Missing**, n* | 1204 | 113 | 297 | 197 | 0 | 161 |
| **Ethnicity** | ***Hispanic**, n (%)* | 1,947 (8.0) | 147 (7.5) | 472 (9.5) | 313 (9.0) | 0 (0) | 372 (11.4) |
| | ***Non-Hispanic**, n (%)* | 22,511 (92.0) | 1,805 (92.5) | 4,486 (90.5) | 3,178 (91.0) | 944 (100) | 2,884 (88.6) |
| | ***Missing**, n* | 672 | 53 | 172 | 117 | 0 | 54 |
| **Patient Status** | ***Outpatient**, n (%)* | 15,569 (62.4) | 1,273 (64.0) | 3,511 (68.7) | 2,584 (71.9) | 524 (55.5) | 0 (0) |
| | ***Inpatient**, n (%)* | 7,807 (31.3) | 591 (29.7) | 1,296 (25.4) | 813 (22.6) | 420 (44.5) | 0 (0) |
| | ***Observation**, n (%)* | 1,519 (6.1) | 119 (6.0) | 291 (5.7) | 189 (5.3) | 0 (0) | 0 (0) |
| | ***Emergency**, n (%)* | 58 (0.2) | 5 (0.3) | 11 (0.2) | 7 (0.2) | 0 (0) | 3,310 (100) |
| | ***Missing**, n* | 177 | 17 | 21 | 15 | 0 | 0 |
| **Hypertension** | ***Yes**, n (%)* | 18,535 (73.8) | 1,447 (72.2) | 3,577 (69.7) | 2,552 (70.7) | 372 (39.4) | 2,323 (70.2) |
| | ***Missing**, n* | 0 | 0 | 0 | 0 | 0 | 0 |
| **Diabetes** | ***Yes**, n (%)* | 6,774 (27.0) | 492 (24.5) | 1,243 (24.2) | 866 (24.0) | 119 (12.6) | 1,001 (30.2) |
| | ***Missing**, n* | 0 | 0 | 0 | 0 | 0 | 0 |
| **Dyslipidemia** | ***Yes**, n (%)* | 5,367 (21.4) | 387 (19.3) | 1,164 (22.7) | 865 (24.0) | 239 (25.3) | 493 (14.9) |
| | ***Missing**, n* | 0 | 0 | 0 | 0 | 0 | 0 |
| **Chronic Kidney Disease** | ***Yes**, n (%)* | 2,394 (9.5) | 182 (9.1) | 350 (6.8) | 251 (7.0) | 108 (11.4) | 442 (13.4) |
| | ***Missing**, n* | 0 | 0 | 0 | 0 | 0 | 0 |
| **Coronary Artery Disease** | ***Yes**, n (%)* | 5,125 (20.4) | 395 (19.7) | 824 (16.1) | 578 (16.0) | 91 (9.6) | 1,103 (33.3) |
| | ***Missing**, n* | 0 | 0 | 0 | 0 | 0 | 0 |

| | | | | | | | |
|---|---|---|---|---|---|---|---|
| **Aortic Root Diameter (mm)** | ***Present**, median (IQR)* | 33.0 (30.0-36.0) | 33.0 (30.0-36.3) | 33.0 (30.0-36.6) | 33.0 (30.0-37.0) | 32.0 (30.0-34.2) | 33.0 (30.0-36.0) |
| | ***Missing**, n* | 2,041 | 165 | 162 | 128 | 1 | 872 |
| **AV Peak Velocity (m/s)** | ***Present**, median (IQR)* | 1.5 (1.3-2.1) | 1.5 (1.2-2.0) | 1.4 (1.2-1.8) | 1.5 (1.2-1.9) | 1.3 (1.2-1.6) | 1.4 (1.2-1.7) |
| | ***Missing**, n* | 2,644 | 205 | 631 | 367 | 10 | 876 |
| **AV Regurgitation** | ***None-Trace**, n (%)* | 16,975 (75.5) | 1,363 (75.3) | 3,676 (77.9) | 2,491 (75.5) | 773 (81.9) | 2,042 (78.4) |
| | ***Mild**, n (%)* | 3,980 (17.7) | 329 (18.2) | 773 (16.4) | 591 (17.9) | 140 (14.8) | 398 (15.3) |
| | *≥ **Moderate**, n (%)* | 1,526 (6.8) | 119 (6.6) | 272 (5.8) | 218 (6.6) | 31 (3.3) | 164 (6.3) |
| | ***Missing**, n* | 2,649 | 194 | 409 | 308 | 0 | 706 |
| **AV Stenosis** | ***None**, n (%)* | 17,118 (79.5) | 1,422 (81.8) | 4,214 (89.6) | 2,907 (88.5) | 850 (90.0) | 2,109 (91.3) |
| | ***Mild-Moderate**, n (%)* | 2,616 (12.1) | 192 (11.0) | 401 (8.5) | 309 (9.4) | 9 (1.0) | 164 (7.1) |
| | ***Severe**, n (%)* | 1,809 (8.4) | 125 (7.2) | 86 (1.8) | 69 (2.1) | 85 (9.0) | 38 (1.6) |
| | ***Missing**, n* | 3,587 | 266 | 429 | 323 | 0 | 999 |
| **AV Structure** | ***Bicuspid**, n (%)* | 207 (1.0) | 5 (0.3) | 52 (1.1) | 37 (1.1) | 8 (0.8) | 10 (0.4) |
| | ***Missing**, n* | 4,053 | 296 | 571 | 389 | 0 | 743 |
| **LV Ejection Fraction (%)** | ***Present**, median (IQR)* | 62.0 (56.1-66.9) | 62.5 (57.0-66.7) | 61.4 (57.0-65.0) | 61.4 (57.0-65.2) | 57.5 (52.2-61.4) | 57.0 (40.0-63.0) |
| | ***Missing**, n* | 942 | 69 | 85 | 58 | 0 | 116 |
| **E\|e' Ratio** | ***Present**, median (IQR)* | 10.4 (7.9-14.3) | 9.9 (7.8-13.7) | 9.2 (7.2-12.1) | 9.3 (7.2-12.3) | 7.0 (5.5-10.6) | 10.6 (7.9-15.0) |
| | ***Missing**, n* | 6,085 | 462 | 781 | 599 | 216 | 1,455 |
| **Global Longitudinal Strain (%)** | ***Present**, median (IQR)* | 18.0 (14.0-20.0)* | 18.0 (15.0-20.0)* | 19.0 (18.0-21.0)* | 19.0 (17.0-21.0)* | 17.6 (13.3-20.7)* | 16.0 (11.0-19.0)* |
| | ***Missing**, n* | 23,663 | 1,888 | 4,669 | 3,297 | 78 | 3,137 |
| **IVSd (mm)** | ***Present**, median (IQR)* | 10.9 (9.1-12.6) | 10.8 (9.0-12.5) | 10.0 (8.8-11.5) | 10.0 (8.9-11.7) | 11.0 (9.0-12.0) | 10.0 (8.9-11.7) |
| | ***Missing**, n* | 1,178 | 82 | 85 | 64 | 0 | 611 |
| **LAIDs (mm)** | ***Present**, median (IQR)* | 40.0 (35.0-46.0) | 40.0 (34.7-45.0) | 39.6 (35.0-45.0) | 40.0 (35.0-45.5) | 40.0 (34.0-46.0) | 41.0 (36.0-48.0) |
| | ***Missing**, n* | 2,861 | 244 | 298 | 220 | 343 | 940 |
| **Increased LA Size** | ***None**, n (%)* | 15,595 (62.1) | 1,251 (62.4) | 3,527 (68.8) | 2,440 (67.6) | 447 (52.9) | 2,166 (65.4) |
| | ***Mild**, n (%)* | 4,543 (18.1) | 354 (17.7) | 805 (15.7) | 570 (15.8) | 125 (14.8) | 403 (12.2) |
| | *≥ **Moderate**, n (%)* | 4,992 (19.9) | 400 (20.0) | 798 (15.6) | 598 (16.6) | 273 (32.3) | 741 (22.4) |
| | ***Missing**, n* | 0 | 0 | 0 | 0 | 99 | 0 |
| **LA Volume ($cm^3$)** | ***Present**, median (IQR)* | 58.5 (45.0-74.4) | 58.5 (45.0-74.0) | 55.2 (43.0-70.0) | 56.0 (43.9-71.0) | 64.5 (46.2-91.3) | 61.9 (47.0-79.0) |
| | ***Missing**, n* | 7,143 | 554 | 1,023 | 730 | 99 | 1,446 |
| **LV Diastolic Dysfunction** | ***None**, n (%)* | 8,583 (48.4) | 702 (50.1) | 2,524 (62.0) | 1,686 (59.7) | 485 (58.7) | 880 (49.7) |
| | ***Mild**, n (%)* | 6,376 (36.0) | 496 (35.4) | 1,214 (29.8) | 892 (31.6) | 182 (22.0) | 473 (26.7) |
| | *≥ **Moderate**, n (%)* | 2,762 (15.6) | 203 (14.5) | 333 (8.2) | 247 (8.7) | 159 (19.2) | 416 (23.5) |
| | ***Missing**, n* | 7,409 | 604 | 1,059 | 783 | 118 | 1,541 |
| **LV ED Volume ($cm^3$)** | ***Present**, median (IQR)* | 96.9 (72.0-130.0) | 97.4 (72.0-127.0) | 105.1 (80.6-136.9) | 105.0 (80.8-137.0) | 128.9 (104.0-166.1) | 130.0 (98.0-170.0) |
| | ***Missing**, n* | 6,053 | 505 | 1,681 | 1,084 | 0 | 821 |
| **LV ES Volume ($cm^3$)** | ***Present**, median (IQR)* | 36.0 (25.2-53.3) | 36.4 (25.0-51.0) | 40.1 (28.9-56.4) | 40.0 (28.9-56.4) | 55.0 (41.9-78.0) | 57.0 (38.0-92.4) |
| | ***Missing**, n* | 6,243 | 515 | 1,695 | 1,096 | 0 | 858 |

| | | | | | | | |
|---|---|---|---|---|---|---|---|
| **LVIDd (mm)** | ***Present**, median (IQR)* | 45.5 (41.0-50.1) | 45.1 (40.5-50.0) | 46.0 (41.8-50.7) | 46.0 (41.8-50.7) | 49.0 (45.0-55.0) | 47.2 (42.1-52.8) |
| | ***Missing**, n* | 1,170 | 85 | 91 | 69 | 0 | 614 |
| **LVIDs (mm)** | ***Present**, median (IQR)* | 29.5 (25.7-34.1) | 29.4 (25.6-33.7) | 30.0 (26.2-34.0) | 30.0 (26.3-34.0) | 33.0 (28.0-38.0) | 32.2 (27.6-39.0) |
| | ***Missing**, n* | 1,323 | 94 | 102 | 74 | 130 | 636 |
| **Elevated LVOT PG** | ***≥ 20 mmHg**, n (%)* | 249 (1.2) | 25 (1.5) | 24 (0.6) | 22 (0.7) | 0 (0) | 2 (0.1) |
| | ***Missing**, n* | 4,600 | 374 | 776 | 512 | 0 | 877 |
| **LVOT Diameter (mm)** | ***Present**, median (IQR)* | 20.0 (19.0-21.8) | 20.0 (19.0-22.0) | 20.0 (19.0-22.0) | 20.0 (19.0-22.0) | 20.2 (18.9-22.0) | 20.0 (19.0-22.0) |
| | ***Missing**, n* | 12,878 | 1,046 | 1,411 | 1,037 | 330 | 2,019 |
| **LVPWd (mm)** | ***Present**, median (IQR)* | 10.4 (9.0-12.0) | 10.4 (9.0-12.0) | 10.0 (8.7-11.2) | 10.0 (8.8-11.3) | 9.0 (8.0-10.0) | 10.0 (8.9-11.4) |
| | ***Missing**, n* | 1,189 | 85 | 90 | 68 | 0 | 612 |
| **Increased LV Size** | ***None**, n (%)* | 16,249 (86.2) | 1,329 (88.4) | 3,660 (85.8) | 2,527 (84.8) | 774 (82.0) | 1,734 (65.9) |
| | ***Mild**, n (%)* | 1,396 (7.4) | 93 (6.2) | 338 (7.9) | 253 (8.5) | 81 (8.6) | 351 (13.3) |
| | ***≥ Moderate**, n (%)* | 1,201 (6.4) | 82 (5.5) | 270 (6.3) | 201 (6.7) | 89 (9.4) | 547 (20.8) |
| | ***Missing**, n* | 6,284 | 501 | 862 | 627 | 0 | 678 |
| **LV Stroke Volume ($cm^3$)** | ***Present**, median (IQR)* | 57.0 (43.0-74.0) | 57.5 (43.2-75.4) | 62.0 (48.3-79.0) | 62.0 (48.1-79.0) | 69.5 (56.4-84.6) | 64.0 (49.1-80.5) |
| | ***Missing**, n* | 6,129 | 516 | 1,735 | 1,126 | 0 | 833 |
| **LV Systolic Dysfunction** | ***None**, n (%)* | 19,820 (84.5) | 1,607 (85.9) | 4,275 (87.6) | 3,005 (87.4) | 692 (73.3) | 1,929 (61.9) |
| | ***Mild**, n (%)* | 1,782 (7.6) | 124 (6.6) | 319 (6.5) | 223 (6.5) | 130 (13.8) | 430 (13.8) |
| | ***≥ Moderate**, n (%)* | 1,849 (7.9) | 140 (7.5) | 284 (5.8) | 212 (6.2) | 122 (12.9) | 755 (24.2) |
| | ***Missing**, n* | 1,679 | 134 | 252 | 168 | 0 | 196 |
| **LV Wall Motion Abnormalities** | ***Any**, n (%)* | 1,704 (20.6) | 143 (21.4) | 348 (22.6) | 249 (22.7) | 116 (12.3) | 630 (44.5) |
| | ***Missing**, n* | 16,855 | 1,338 | 3,589 | 2,512 | 0 | 1,895 |
| **Increased LV Wall Thickness** | ***Any**, n (%)* | 12,509 (54.8) | 990 (54.0) | 1,971 (40.2) | 1,465 (42.7) | 232 (24.6) | 1,206 (44.1) |
| | ***≥ Moderate**, n (%)* | 3,021 (13.2) | 239 (13.0) | 315 (6.4) | 250 (7.3) | 46 (4.9) | 258 (9.4) |
| | ***Missing**, n* | 2,324 | 172 | 229 | 174 | 0 | 575 |
| **MV Regurgitation** | ***None-Trace**, n (%)* | 12,041 (50.7) | 996 (52.3) | 2,976 (59.4) | 1,967 (56.0) | 479 (57.8) | 1,441 (51.5) |
| | ***Mild**, n (%)* | 7,968 (33.6) | 626 (32.9) | 1,419 (28.3) | 1,062 (30.2) | 209 (25.2) | 815 (29.1) |
| | ***≥ Moderate**, n (%)* | 3,718 (15.7) | 282 (14.8) | 612 (12.2) | 482 (13.7) | 141 (17.0) | 544 (19.4) |
| | ***Missing**, n* | 1,403 | 101 | 123 | 97 | 115 | 510 |
| **MV Stenosis** | ***Any**, n (%)* | 758 (4.1) | 54 (3.7) | 89 (2.1) | 67 (2.3) | 11 (1.3) | 71 (3.4) |
| | ***Missing**, n* | 6,823 | 542 | 928 | 693 | 115 | 1,248 |
| **Pericardial Effusion** | ***Any**, n (%)* | 935 (4.0) | 71 (3.8) | 21 (3.4) | 21 (3.4) | 47 (5.0) | 305 (10.9) |
| | ***Missing**, n* | 1,608 | 121 | 4,507 | 2,989 | 0 | 514 |
| **RA Transverse Dimension (mm)** | ***Present**, median (IQR)* | 37.2 (33.0-43.0) | 38.0 (33.0-43.0) | 37.0 (33.0-42.0) | 38.0 (33.0-43.0) | 40.5 (35.0-45.0) | 40.0 (34.0-46.0) |
| | ***Missing**, n* | 14,163 | 1,147 | 2,233 | 1,541 | 0 | 1,749 |
| **Elevated RA Pressure** | ***≥ 8 mmHg**, n (%)* | 4,596 (21.0) | 344 (19.5) | 722 (15.5) | 485 (14.7) | 531 (56.2) | 1,200 (43.2) |
| | ***Missing**, n* | 3,196 | 239 | 484 | 319 | 0 | 535 |
| **Increased RA Size** | ***Any**, n (%)* | 2,988 (29.8) | 231 (28.2) | 507 (29.3) | 374 (29.5) | 220 (23.3) | 541 (41.3) |

| | | | | | | | |
|---|---|---|---|---|---|---|---|
| | ***Missing**, n* | 15,115 | 1,186 | 3,398 | 2,341 | 0 | 2,000 |
| **RVIDd (mm)** | ***Present**, median (IQR)* | 31.0 (27.1-35.1) | 31.0 (27.2-35.0) | 30.9 (27.0-34.9) | 31.0 (27.0-35.1) | 31.0 (28.0-35.0) | 32.0 (28.2-36.3) |
| | ***Missing**, n* | 9,632 | 791 | 1,700 | 1,196 | 0 | 999 |
| **Increased RV Size** | ***None**, n (%)* | 19,229 (83.8) | 1,551 (85.3) | 4,088 (85.9) | 2,843 (84.6) | 738 (78.2) | 1,955 (70.0) |
| | ***Mild**, n (%)* | 2,313 (10.1) | 170 (9.3) | 457 (9.6) | 359 (10.7) | 94 (10.0) | 427 (15.3) |
| | ***≥ Moderate**, n (%)* | 1,417 (6.2) | 98 (5.4) | 215 (4.5) | 158 (4.7) | 112 (11.9) | 411 (14.7) |
| | ***Missing**, n* | 2,171 | 186 | 370 | 248 | 0 | 517 |
| **RV Systolic Pressure (mmHg)** | ***Present**, median (IQR)* | 29.0 (23.0-38.0) | 28.0 (22.4-37.0) | 27.5 (22.5-35.0) | 27.1 (22.1-34.4) | 28.0 (22.0-38.0) | 37.0 (28.0-49.0) |
| | ***Missing**, n* | 7,923 | 589 | 1,832 | 1,192 | 255 | 1,373 |
| **RV S' (cm/s)** | ***Present**, median (IQR)* | 12.3 (10.4-14.4) | 12.5 (10.9-14.6) | 12.6 (10.9-14.7) | 12.6 (10.9-14.6) | 13.0 (11.0-15.0) | 11.9 (9.5-14.0) |
| | ***Missing**, n* | 10,403 | 864 | 1,499 | 1,075 | 113 | 1,349 |
| **RV Systolic Dysfunction** | ***Any**, n (%)* | 2,589 (11.3) | 196 (10.8) | 371 (7.8) | 263 (7.9) | 76 (8.1) | 819 (29.1) |
| | ***Missing**, n* | 2,243 | 198 | 387 | 261 | 0 | 497 |
| **TAPSE (mm)** | ***Present**, median (IQR)* | 21.0 (17.9-24.9) | 21.5 (18.0-25.0) | 21.2 (18.0-24.8) | 21.1 (18.0-24.8) | 21.0 (17.0-26.0) | 19.0 (16.0-23.0) |
| | ***Missing**, n* | 9,774 | 798 | 1,396 | 971 | 0 | 1,279 |
| **TV Peak Gradient (mmHg)** | ***Present**, median (IQR)* | 25.0 (19.6-32.7) | 24.0 (19.0-31.4) | 24.0 (19.0-30.0) | 23.9 (19.0-30.0) | 22.0 (17.0-30.0) | 29.0 (22.0-40.0) |
| | ***Missing**, n* | 6,493 | 488 | 1,615 | 1,042 | 255 | 1,142 |
| **TV Regurgitation** | ***None-Trace**, n (%)* | 12,301 (50.9) | 1,004 (52.1) | 2,886 (56.9) | 1,941 (54.4) | 532 (56.4) | 1,430 (46.7) |
| | ***Mild**, n (%)* | 8,199 (33.9) | 630 (32.7) | 1,580 (31.1) | 1,173 (32.9) | 310 (32.8) | 939 (30.7) |
| | ***≥ Moderate**, n (%)* | 3,666 (15.2) | 294 (15.2) | 610 (12.0) | 454 (12.7) | 102 (10.8) | 692 (22.6) |
| | ***Missing**, n* | 964 | 77 | 54 | 40 | 0 | 249 |

Demographics and summary statistics of all cohorts used in this study. All summary statistics are computed after excluding missing values and provided at the study level since age and labels are unique to each echocardiography study. * = Global longitudinal strain values are presented as positive to aid readability.
AV = aortic valve; ED = end-diastolic; ES = end-systolic; IVSd = interventricular septum thickness at diastole; IQR = interquartile range; LA = left atrium; LV = left ventricle; LAIDs = left atrial internal diameter at systole; LVIDd = left ventricular internal diameter at diastole; LVIDs = left ventricular internal diameter at systole; LVOT = left ventricular outflow tract; LVPWd = left ventricular posterior wall thickness at diastole; PG = pressure gradient; RA = right atrium; RV = right ventricle; RVIDd = right ventricular internal diameter at diastole; RV S' = right ventricular systolic excursion velocity; TAPSE = tricuspid annular plane systolic excursion; TTE = transthoracic echocardiography; TV = tricuspid valve; YNHHS = Yale-New Haven Health System.

#### eTable 3 | Detailed Multi-task Classification Performance Evaluation

| | | Cohort | AUC | AP | Sensitivity | Specificity | Brier Score | n/N |
|---|---|---|---|---|---|---|---|---|
| **AV Regurgitation** | *None-Trace* | YNHHS | 0.86 (0.85, 0.87) | 0.95 (0.95, 0.96) | 0.76 (0.75, 0.77) | 0.81 (0.79, 0.83) | 0.12 (0.12, 0.13) | 3,676/4,721 |
| | | RVENet+ | 0.83 (0.80, 0.85) | 0.95 (0.93, 0.96) | 0.72 (0.69, 0.74) | 0.79 (0.73, 0.84) | 0.12 (0.11, 0.13) | 773/944 |
| | *Mild* | YNHHS | 0.79 (0.78, 0.80) | 0.40 (0.37, 0.43) | 0.84 (0.82, 0.86) | 0.60 (0.59, 0.61) | 0.12 (0.11, 0.12) | 773/4,721 |
| | | RVENet+ | 0.79 (0.75, 0.82) | 0.40 (0.34, 0.48) | 0.81 (0.76, 0.87) | 0.62 (0.59, 0.65) | 0.11 (0.10, 0.12) | 140/944 |
| | *≥ Moderate* | YNHHS | 0.92 (0.91, 0.93) | 0.52 (0.47, 0.57) | 0.85 (0.82, 0.89) | 0.83 (0.82, 0.84) | 0.04 (0.04, 0.05) | 272/4,721 |
| | | RVENet+ | 0.85 (0.81, 0.89) | 0.15 (0.10, 0.25) | 0.77 (0.64, 0.90) | 0.76 (0.74, 0.78) | 0.03 (0.02, 0.04) | 31/944 |
| | *Mean* | YNHHS | 0.86 (0.85, 0.87) | 0.62 (0.61, 0.64) | 0.82 (0.80, 0.83) | 0.75 (0.74, 0.75) | 0.10 (0.09, 0.10) | N/A |
| | | RVENet+ | 0.82 (0.80, 0.85) | 0.50 (0.48, 0.54) | 0.77 (0.72, 0.82) | 0.72 (0.70, 0.74) | 0.09 (0.08, 0.09) | N/A |
| **AV Stenosis** | *None* | YNHHS | 0.95 (0.94, 0.96) | 0.99 (0.99, 0.99) | 0.91 (0.91, 0.92) | 0.85 (0.82, 0.88) | 0.05 (0.05, 0.05) | 4,214/4,701 |
| | | RVENet+ | 0.98 (0.96, 0.99) | 1.00 (0.99, 1.00) | 0.88 (0.87, 0.90) | 0.95 (0.90, 0.98) | 0.03 (0.03, 0.04) | 850/944 |
| | *Mild-Moderate* | YNHHS | 0.93 (0.92, 0.94) | 0.54 (0.50, 0.59) | 0.82 (0.79, 0.85) | 0.90 (0.89, 0.90) | 0.06 (0.05, 0.06) | 401/4,701 |
| | | RVENet+ | 0.70 (0.57, 0.81) | 0.02 (0.01, 0.04) | 0.44 (0.14, 0.73) | 0.80 (0.78, 0.82) | 0.03 (0.02, 0.03) | 9/944 |
| | *Severe* | YNHHS | 0.98 (0.98, 0.99) | 0.58 (0.48, 0.67) | 0.94 (0.89, 0.98) | 0.95 (0.94, 0.95) | 0.01 (0.01, 0.01) | 86/4,701 |
| | | RVENet+ | 1.00 (0.99, 1.00) | 0.97 (0.96, 0.99) | 0.96 (0.93, 0.99) | 0.98 (0.97, 0.99) | 0.04 (0.04, 0.05) | 85/944 |
| | *Mean* | YNHHS | 0.96 (0.95, 0.96) | 0.70 (0.67, 0.74) | 0.89 (0.87, 0.91) | 0.90 (0.89, 0.91) | 0.04 (0.04, 0.04) | N/A |
| | | RVENet+ | 0.89 (0.85, 0.93) | 0.66 (0.66, 0.67) | 0.76 (0.66, 0.86) | 0.91 (0.89, 0.92) | 0.03 (0.03, 0.04) | N/A |
| **AV Structure** | *Bicuspid* | YNHHS | 0.81 (0.75, 0.86) | 1.00 (1.00, 1.00) | 0.70 (0.69, 0.71) | 0.69 (0.58, 0.80) | 0.01 (0.01, 0.01) | 52/4,559 |
| | | RVENet+ | 0.83 (0.65, 0.96) | 0.14 (0.03, 0.39) | 0.88 (0.67, 1.00) | 0.60 (0.58, 0.63) | 0.01 (0.00, 0.01) | 8/944 |
| **Increased LA Size** | *Mild* | YNHHS | 0.75 (0.74, 0.77) | 0.36 (0.33, 0.38) | 0.67 (0.65, 0.70) | 0.70 (0.69, 0.72) | 0.12 (0.11, 0.12) | 805/5,130 |
| | | RVENet+ | 0.65 (0.60, 0.69) | 0.23 (0.19, 0.29) | 0.52 (0.44, 0.60) | 0.64 (0.62, 0.67) | 0.13 (0.12, 0.14) | 125/845 |
| | *≥ Moderate* | YNHHS | 0.89 (0.88, 0.90) | 0.59 (0.56, 0.63) | 0.79 (0.76, 0.81) | 0.83 (0.82, 0.84) | 0.09 (0.09, 0.09) | 798/5,130 |
| | | RVENet+ | 0.90 (0.88, 0.92) | 0.77 (0.73, 0.83) | 0.78 (0.74, 0.82) | 0.88 (0.86, 0.91) | 0.16 (0.15, 0.17) | 273/845 |
| | *None* | YNHHS | 0.84 (0.83, 0.85) | 0.92 (0.91, 0.92) | 0.74 (0.73, 0.75) | 0.78 (0.76, 0.79) | 0.15 (0.15, 0.16) | 3,527/5,130 |
| | | RVENet+ | 0.87 (0.84, 0.89) | 0.88 (0.85, 0.90) | 0.82 (0.79, 0.85) | 0.79 (0.76, 0.82) | 0.16 (0.15, 0.18) | 447/845 |
| | *Mean* | YNHHS | 0.83 (0.82, 0.83) | 0.62 (0.61, 0.64) | 0.73 (0.72, 0.75) | 0.77 (0.76, 0.78) | 0.12 (0.12, 0.12) | N/A |
| | | RVENet+ | 0.81 (0.78, 0.83) | 0.63 (0.61, 0.66) | 0.70 (0.67, 0.73) | 0.77 (0.76, 0.79) | 0.15 (0.14, 0.16) | N/A |
| **LV Diastolic Dysfunction** | *Mild* | YNHHS | 0.80 (0.79, 0.81) | 0.63 (0.60, 0.66) | 0.59 (0.57, 0.61) | 0.81 (0.80, 0.83) | 0.16 (0.16, 0.16) | 1,214/4,071 |
| | | RVENet+ | 0.79 (0.76, 0.82) | 0.52 (0.46, 0.58) | 0.47 (0.41, 0.53) | 0.87 (0.85, 0.89) | 0.14 (0.13, 0.15) | 182/826 |
| | *≥ Moderate* | YNHHS | 0.92 (0.91, 0.93) | 0.52 (0.48, 0.57) | 0.88 (0.85, 0.91) | 0.81 (0.80, 0.82) | 0.05 (0.05, 0.06) | 333/4,071 |
| | | RVENet+ | 0.92 (0.90, 0.93) | 0.67 (0.60, 0.74) | 0.95 (0.92, 0.98) | 0.78 (0.76, 0.81) | 0.10 (0.09, 0.11) | 159/826 |
| | *None* | YNHHS | 0.84 (0.83, 0.85) | 0.90 (0.89, 0.91) | 0.81 (0.79, 0.82) | 0.69 (0.67, 0.71) | 0.16 (0.15, 0.16) | 2,524/4,071 |
| | | RVENet+ | 0.94 (0.92, 0.95) | 0.95 (0.93, 0.96) | 0.91 (0.89, 0.93) | 0.81 (0.77, 0.84) | 0.10 (0.09, 0.11) | 485/826 |
| | *Mean* | YNHHS | 0.85 (0.85, 0.86) | 0.68 (0.67, 0.70) | 0.76 (0.75, 0.77) | 0.77 (0.76, 0.78) | 0.12 (0.12, 0.13) | N/A |
| | | RVENet+ | 0.88 (0.87, 0.90) | 0.71 (0.68, 0.74) | 0.78 (0.75, 0.80) | 0.82 (0.80, 0.83) | 0.11 (0.11, 0.12) | N/A |
| **Elevated LVOT PG** | *≥ 20 mmHg* | YNHHS | 0.94 (0.88, 0.99) | 0.49 (0.33, 0.69) | 0.88 (0.75, 1.00) | 0.95 (0.95, 0.96) | 0.00 (0.00, 0.01) | 24/4,354 |
| | | RVENet+ | N/A | N/A | N/A | N/A | N/A | 0/944 |

| | | | | | | | | |
|---|---|---|---|---|---|---|---|---|
| **Increased LV Size** | *Mild* | YNHHS | 0.83 (0.82, 0.85) | 0.30 (0.26, 0.34) | 0.71 (0.66, 0.75) | 0.80 (0.79, 0.81) | 0.06 (0.06, 0.07) | 338/4,268 |
| | | RVENet+ | 0.78 (0.73, 0.82) | 0.26 (0.20, 0.35) | 0.58 (0.49, 0.67) | 0.80 (0.77, 0.82) | 0.07 (0.06, 0.08) | 81/944 |
| | | EchoNet-LVH | 0.84 (0.84, 0.85) | 0.51 (0.49, 0.53) | 0.56 (0.55, 0.58) | 0.89 (0.88, 0.89) | 0.14 (0.14, 0.15) | 2,174/11,713 |
| | *≥ Moderate* | YNHHS | 0.94 (0.93, 0.95) | 0.65 (0.60, 0.70) | 0.81 (0.77, 0.85) | 0.92 (0.91, 0.93) | 0.04 (0.03, 0.04) | 270/4,268 |
| | | RVENet+ | 0.91 (0.88, 0.93) | 0.60 (0.52, 0.68) | 0.71 (0.63, 0.78) | 0.90 (0.88, 0.92) | 0.06 (0.05, 0.07) | 89/944 |
| | | EchoNet-LVH | 0.98 (0.98, 0.99) | 0.73 (0.69, 0.76) | 0.95 (0.93, 0.97) | 0.93 (0.92, 0.93) | 0.02 (0.02, 0.02) | 418/11,713 |
| | *None* | YNHHS | 0.91 (0.90, 0.92) | 0.98 (0.98, 0.98) | 0.89 (0.88, 0.90) | 0.76 (0.73, 0.79) | 0.08 (0.07, 0.08) | 3,660/4,268 |
| | | RVENet+ | 0.88 (0.85, 0.90) | 0.96 (0.95, 0.97) | 0.88 (0.86, 0.90) | 0.65 (0.59, 0.71) | 0.10 (0.09, 0.11) | 774/944 |
| | | EchoNet-LVH | 0.89 (0.88, 0.89) | 0.96 (0.96, 0.97) | 0.94 (0.94, 0.94) | 0.56 (0.55, 0.58) | 0.13 (0.12, 0.13) | 9,121/11,713 |
| | *Mean* | YNHHS | 0.89 (0.88, 0.90) | 0.64 (0.63, 0.66) | 0.80 (0.78, 0.82) | 0.83 (0.82, 0.84) | 0.06 (0.06, 0.06) | N/A |
| | | RVENet+ | 0.85 (0.83, 0.88) | 0.61 (0.58, 0.64) | 0.72 (0.68, 0.76) | 0.78 (0.77, 0.80) | 0.08 (0.07, 0.09) | N/A |
| | | EchoNet-LVH | 0.91 (0.90, 0.91) | 0.73 (0.72, 0.75) | 0.82 (0.81, 0.83) | 0.79 (0.79, 0.80) | 0.09 (0.09, 0.10) | N/A |
| **LV Systolic Dysfunction** | *Mild* | YNHHS | 0.92 (0.91, 0.93) | 0.41 (0.37, 0.45) | 0.89 (0.86, 0.92) | 0.84 (0.83, 0.85) | 0.05 (0.04, 0.05) | 319/4,878 |
| | | RVENet+ | 0.71 (0.67, 0.75) | 0.29 (0.24, 0.36) | 0.50 (0.42, 0.57) | 0.76 (0.74, 0.79) | 0.11 (0.10, 0.13) | 130/944 |
| | | EchoNet-Dynamic | 0.78 (0.77, 0.79) | 0.35 (0.33, 0.37) | 0.58 (0.56, 0.60) | 0.80 (0.79, 0.81) | 0.12 (0.12, 0.12) | 1,533/10,032 |
| | *≥ Moderate* | YNHHS | 0.98 (0.98, 0.99) | 0.85 (0.82, 0.88) | 0.92 (0.90, 0.95) | 0.94 (0.94, 0.95) | 0.02 (0.02, 0.02) | 284/4,878 |
| | | RVENet+ | 0.99 (0.98, 0.99) | 0.94 (0.91, 0.96) | 0.97 (0.94, 0.99) | 0.89 (0.87, 0.91) | 0.03 (0.03, 0.04) | 122/944 |
| | | EchoNet-Dynamic | 0.94 (0.94, 0.95) | 0.78 (0.76, 0.79) | 0.80 (0.78, 0.82) | 0.91 (0.91, 0.92) | 0.06 (0.05, 0.06) | 1,264/10,032 |
| | *None* | YNHHS | 0.97 (0.96, 0.97) | 0.99 (0.99, 1.00) | 0.91 (0.90, 0.92) | 0.91 (0.89, 0.93) | 0.04 (0.04, 0.05) | 4,275/4,878 |
| | | RVENet+ | 0.88 (0.85, 0.90) | 0.93 (0.91, 0.95) | 0.85 (0.83, 0.88) | 0.75 (0.70, 0.79) | 0.11 (0.10, 0.12) | 692/944 |
| | | EchoNet-Dynamic | 0.89 (0.88, 0.90) | 0.95 (0.94, 0.95) | 0.90 (0.89, 0.90) | 0.71 (0.69, 0.72) | 0.12 (0.12, 0.13) | 7,235/10,032 |
| | *Mean* | YNHHS | 0.96 (0.95, 0.96) | 0.75 (0.73, 0.77) | 0.91 (0.90, 0.92) | 0.90 (0.89, 0.91) | 0.04 (0.03, 0.04) | N/A |
| | | RVENet+ | 0.86 (0.84, 0.88) | 0.72 (0.70, 0.75) | 0.77 (0.74, 0.80) | 0.80 (0.78, 0.82) | 0.09 (0.08, 0.09) | N/A |
| | | EchoNet-Dynamic | 0.87 (0.86, 0.88) | 0.69 (0.68, 0.70) | 0.76 (0.75, 0.77) | 0.81 (0.80, 0.81) | 0.10 (0.10, 0.10) | N/A |
| **LV Wall Motion Abnormalities** | *Any* | YNHHS | 0.88 (0.86, 0.90) | 0.75 (0.71, 0.78) | 0.76 (0.73, 0.80) | 0.85 (0.83, 0.86) | 0.11 (0.10, 0.12) | 348/1,541 |
| | | RVENet+ | 0.98 (0.97, 0.99) | 0.91 (0.88, 0.94) | 0.98 (0.96, 1.00) | 0.71 (0.69, 0.74) | 0.06 (0.06, 0.07) | 116/944 |
| **Increased LV Wall Thickness** | *Any* | YNHHS | 0.85 (0.84, 0.86) | 0.89 (0.88, 0.90) | 0.82 (0.81, 0.83) | 0.69 (0.67, 0.70) | 0.16 (0.15, 0.16) | 1,971/4,901 |
| | | RVENet+ | 0.83 (0.80, 0.85) | 0.63 (0.57, 0.68) | 0.50 (0.45, 0.56) | 0.90 (0.88, 0.92) | 0.14 (0.13, 0.15) | 232/944 |
| | | EchoNet-LVH | 0.84 (0.84, 0.85) | 0.94 (0.94, 0.94) | 0.84 (0.84, 0.85) | 0.63 (0.62, 0.65) | 0.14 (0.14, 0.14) | 2,971/11,651 |
| | *≥ Moderate* | YNHHS | 0.91 (0.90, 0.93) | 0.99 (0.99, 0.99) | 0.90 (0.89, 0.90) | 0.75 (0.71, 0.79) | 0.04 (0.04, 0.04) | 315/4,815 |

| | | | | | | | | |
|---|---|---|---|---|---|---|---|---|
| | | RVENet+ | 0.91 (0.88, 0.94) | 0.39 (0.30, 0.54) | 0.50 (0.38, 0.62) | 0.96 (0.95, 0.97) | 0.04 (0.03, 0.05) | 46/944 |
| | | EchoNet-LVH | 0.89 (0.88, 0.90) | 0.99 (0.99, 0.99) | 0.92 (0.92, 0.93) | 0.57 (0.54, 0.60) | 0.04 (0.04, 0.05) | 703/11,651 |
| **MV Regurgitation** | *None-Trace* | YNHHS | 0.84 (0.84, 0.85) | 0.88 (0.87, 0.89) | 0.85 (0.84, 0.86) | 0.66 (0.64, 0.67) | 0.16 (0.15, 0.16) | 2,976/5,007 |
| | | RVENet+ | 0.91 (0.90, 0.93) | 0.91 (0.88, 0.94) | 0.92 (0.90, 0.94) | 0.73 (0.69, 0.77) | 0.12 (0.11, 0.13) | 479/829 |
| | *Mild* | YNHHS | 0.73 (0.72, 0.75) | 0.49 (0.47, 0.51) | 0.70 (0.68, 0.72) | 0.65 (0.63, 0.66) | 0.18 (0.17, 0.18) | 1,419/5,007 |
| | | RVENet+ | 0.78 (0.75, 0.81) | 0.52 (0.46, 0.58) | 0.63 (0.57, 0.68) | 0.78 (0.75, 0.81) | 0.16 (0.15, 0.17) | 209/829 |
| | *≥ Moderate* | YNHHS | 0.91 (0.90, 0.91) | 0.60 (0.57, 0.63) | 0.74 (0.71, 0.77) | 0.87 (0.86, 0.88) | 0.07 (0.07, 0.08) | 612/5,007 |
| | | RVENet+ | 0.92 (0.91, 0.94) | 0.67 (0.61, 0.74) | 0.88 (0.83, 0.92) | 0.83 (0.80, 0.85) | 0.09 (0.08, 0.09) | 141/829 |
| | *Mean* | YNHHS | 0.83 (0.82, 0.84) | 0.66 (0.64, 0.67) | 0.76 (0.75, 0.77) | 0.72 (0.72, 0.73) | 0.14 (0.13, 0.14) | N/A |
| | | RVENet+ | 0.87 (0.86, 0.89) | 0.70 (0.67, 0.74) | 0.81 (0.79, 0.83) | 0.78 (0.76, 0.79) | 0.12 (0.12, 0.13) | N/A |
| **MV Stenosis** | *Any* | YNHHS | 0.96 (0.94, 0.98) | 1.00 (1.00, 1.00) | 0.96 (0.96, 0.97) | 0.82 (0.75, 0.88) | 0.01 (0.01, 0.02) | 89/4,202 |
| | | RVENet+ | 1.00 (1.00, 1.00) | 0.95 (0.85, 1.00) | 1.00 (1.00, 1.00) | 0.95 (0.94, 0.97) | 0.01 (0.00, 0.01) | 11/829 |
| **Pericardial Effusion** | *Any* | YNHHS | 0.91 (0.83, 0.97) | 0.99 (0.98, 1.00) | 0.91 (0.89, 0.93) | 0.86 (0.73, 0.96) | 0.02 (0.02, 0.03) | 21/623 |
| | | RVENet+ | 0.94 (0.89, 0.98) | 0.86 (0.78, 0.93) | 0.68 (0.56, 0.79) | 1.00 (1.00, 1.00) | 0.04 (0.03, 0.05) | 47/944 |
| **Elevated RA Pressure** | *≥ 8 mmHg* | YNHHS | 0.81 (0.80, 0.83) | 0.53 (0.50, 0.56) | 0.62 (0.59, 0.65) | 0.82 (0.81, 0.83) | 0.10 (0.10, 0.11) | 722/4,646 |
| | | RVENet+ | 0.80 (0.78, 0.82) | 0.86 (0.84, 0.88) | 0.49 (0.46, 0.53) | 0.96 (0.94, 0.98) | 0.36 (0.34, 0.37) | 531/944 |
| **Increased RA Size** | *Any* | YNHHS | 0.77 (0.75, 0.79) | 0.88 (0.86, 0.89) | 0.71 (0.69, 0.73) | 0.69 (0.66, 0.72) | 0.17 (0.16, 0.17) | 507/1,732 |
| | | RVENet+ | 0.75 (0.72, 0.78) | 0.50 (0.44, 0.57) | 0.70 (0.65, 0.76) | 0.68 (0.65, 0.71) | 0.15 (0.14, 0.16) | 220/944 |
| **Increased RV Size** | *Mild* | YNHHS | 0.78 (0.76, 0.79) | 0.24 (0.22, 0.27) | 0.60 (0.56, 0.63) | 0.77 (0.76, 0.78) | 0.08 (0.07, 0.08) | 457/4,760 |
| | | RVENet+ | 0.64 (0.60, 0.69) | 0.15 (0.12, 0.20) | 0.40 (0.33, 0.49) | 0.74 (0.72, 0.77) | 0.09 (0.08, 0.10) | 94/944 |
| | *≥ Moderate* | YNHHS | 0.88 (0.86, 0.90) | 0.34 (0.29, 0.40) | 0.72 (0.66, 0.77) | 0.85 (0.84, 0.86) | 0.04 (0.03, 0.04) | 215/4,760 |
| | | RVENet+ | 0.83 (0.79, 0.86) | 0.48 (0.41, 0.57) | 0.64 (0.56, 0.72) | 0.81 (0.79, 0.84) | 0.09 (0.08, 0.11) | 112/944 |
| | *None* | YNHHS | 0.84 (0.82, 0.85) | 0.97 (0.96, 0.97) | 0.83 (0.82, 0.84) | 0.64 (0.62, 0.68) | 0.10 (0.09, 0.10) | 4,088/4,760 |
| | | RVENet+ | 0.78 (0.75, 0.81) | 0.92 (0.91, 0.94) | 0.82 (0.80, 0.85) | 0.57 (0.52, 0.63) | 0.15 (0.13, 0.16) | 738/944 |
| | *Mean* | YNHHS | 0.83 (0.82, 0.84) | 0.52 (0.50, 0.54) | 0.72 (0.69, 0.74) | 0.75 (0.74, 0.76) | 0.07 (0.07, 0.07) | N/A |
| | | RVENet+ | 0.75 (0.73, 0.78) | 0.52 (0.49, 0.55) | 0.62 (0.59, 0.66) | 0.71 (0.69, 0.73) | 0.11 (0.10, 0.12) | N/A |
| **RV Systolic Dysfunction** | *Any* | YNHHS | 0.93 (0.91, 0.94) | 0.99 (0.99, 0.99) | 0.89 (0.88, 0.90) | 0.80 (0.77, 0.84) | 0.05 (0.04, 0.05) | 371/4,743 |
| | | RVENet+ | 0.94 (0.92, 0.96) | 0.64 (0.56, 0.73) | 0.92 (0.87, 0.97) | 0.82 (0.80, 0.84) | 0.05 (0.04, 0.05) | 76/944 |
| **TV Regurgitation** | *None-Trace* | YNHHS | 0.83 (0.82, 0.84) | 0.85 (0.84, 0.86) | 0.82 (0.80, 0.83) | 0.67 (0.65, 0.68) | 0.17 (0.16, 0.17) | 2,886/5,076 |
| | | RVENet+ | 0.86 (0.84, 0.88) | 0.88 (0.85, 0.90) | 0.86 (0.84, 0.89) | 0.71 (0.68, 0.75) | 0.16 (0.15, 0.17) | 532/944 |
| | *Mild* | YNHHS | 0.72 (0.71, 0.73) | 0.50 (0.48, 0.52) | 0.64 (0.62, 0.66) | 0.68 (0.67, 0.69) | 0.19 (0.18, 0.19) | 1,580/5,076 |
| | | RVENet+ | 0.75 (0.72, 0.77) | 0.55 (0.51, 0.61) | 0.62 (0.58, 0.67) | 0.76 (0.73, 0.79) | 0.20 (0.19, 0.21) | 310/944 |
| | *≥ Moderate* | YNHHS | 0.90 (0.89, 0.91) | 0.61 (0.58, 0.65) | 0.76 (0.73, 0.78) | 0.86 (0.86, 0.87) | 0.07 (0.07, 0.08) | 610/5,076 |
| | | RVENet+ | 0.91 (0.90, 0.93) | 0.58 (0.49, 0.65) | 0.86 (0.81, 0.92) | 0.80 (0.77, 0.82) | 0.07 (0.06, 0.07) | 102/944 |
| | *Mean* | YNHHS | 0.82 (0.81, 0.82) | 0.66 (0.64, 0.67) | 0.74 (0.73, 0.75) | 0.74 (0.73, 0.74) | 0.14 (0.14, 0.15) | N/A |
| | | RVENet+ | 0.84 (0.83, 0.86) | 0.67 (0.64, 0.70) | 0.78 (0.76, 0.81) | 0.76 (0.74, 0.77) | 0.14 (0.14, 0.15) | N/A |

Multi-task validation of PanEcho on diagnostic classification tasks. Results are presented on the internal YNHHS validation cohort and external RVENet+, EchoNet-Dynamic, and EchoNet-LVH cohorts. Values in parentheses represent bootstrapped 95% confidence intervals. For multi-class classification tasks, metrics are presented for each individual class, followed by the "Mean" overall performance across classes. n = number of studies meeting the label definition; N = number of non-missing labels; * = moderate or higher; ^ = severe.
AP = average precision; AUC = area under the receiver operator characteristic curve; AV = aortic valve; LA = left atrium; LV = left ventricle; LVOT = left ventricular outflow tract; PG = pressure gradient; RA = right atrium; RV = right ventricle; TV = tricuspid valve; YNHHS = Yale-New Haven Health System.

**eTable 4 | Detailed Multi-task Regression Performance Evaluation**

| | | MAE | MAD | RMSE | NMAE | r | Median (IQR) | N |
|---|---|---|---|---|---|---|---|---|
| **Aortic Root Dimension (mm)** | YNHHS | 2.82 (2.77, 2.87) | 2.33 (2.26, 2.39) | 3.62 (3.55, 3.69) | 0.08 (0.08, 0.09) | 0.69 (0.68, 0.70) | 3.3 (3.0-3.7) | 4,968 |
| | RVENet+ | 2.16 (2.08, 2.26) | 1.83 (1.72, 1.94) | 2.72 (2.61, 2.83) | 0.07 (0.07, 0.07) | 0.51 (0.47, 0.55) | 3.2 (3.0-3.4) | 943 |
| **AV Peak Velocity (m/s)** | YNHHS | 0.31 (0.30, 0.32) | 0.24 (0.23, 0.24) | 0.43 (0.42, 0.44) | 0.19 (0.18, 0.19) | 0.79 (0.78, 0.81) | 1.4 (1.2-1.8) | 4,499 |
| | RVENet+ | 0.36 (0.34, 0.38) | 0.25 (0.23, 0.26) | 0.51 (0.48, 0.54) | 0.23 (0.22, 0.24) | 0.84 (0.81, 0.86) | 1.3 (1.2-1.6) | 934 |
| **LV Ejection Fraction (%)** | YNHHS | 4.17 (4.08, 4.25) | 3.21 (3.13, 3.28) | 5.55 (5.43, 5.68) | 0.07 (0.07, 0.07) | 0.85 (0.84, 0.86) | 61.7 (57.0-65.1) | 5,045 |
| | RVENet+ | 4.53 (4.33, 4.71) | 3.61 (3.42, 3.83) | 5.83 (5.55, 6.08) | 0.08 (0.08, 0.09) | 0.87 (0.86, 0.89) | 57.5 (52.2-61.4) | 944 |
| | EchoNet-Dynamic | 5.53 (5.45, 5.62) | 4.02 (3.93, 4.10) | 7.70 (7.57, 7.84) | 0.10 (0.10, 0.10) | 0.79 (0.78, 0.80) | 59.2 (51.6-64.0) | 10,032 |
| **E\|e' Ratio** | YNHHS | 1.97 (1.92, 2.01) | 1.54 (1.50, 1.58) | 2.62 (2.56, 2.69) | 0.19 (0.19, 0.20) | 0.80 (0.79, 0.81) | 9.2 (7.2-12.1) | 4,349 |
| | RVENet+ | 2.18 (2.07, 2.31) | 1.61 (1.51, 1.72) | 2.95 (2.76, 3.12) | 0.24 (0.23, 0.26) | 0.84 (0.82, 0.86) | 7.0 (5.5-10.6) | 728 |
| **Global Longitudinal Strain (%)** | YNHHS | 1.89 (1.78, 2.01) | 1.56 (1.39, 1.67) | 2.44 (2.28, 2.60) | 0.10 (0.09, 0.11) | 0.65 (0.59, 0.71) | 19.0 (18.0-21.0)* | 461 |
| | RVENet+ | 2.65 (2.54, 2.75) | 2.28 (2.16, 2.40) | 3.28 (3.15, 3.41) | 0.16 (0.15, 0.17) | 0.82 (0.80, 0.84) | 17.6 (13.3-20.7) | 866 |
| **IVSd (mm)** | YNHHS | 1.30 (1.27, 1.33) | 1.02 (1.00, 1.05) | 1.79 (1.71, 1.90) | 0.13 (0.12, 0.13) | 0.70 (0.68, 0.72) | 1.0 (0.9-1.1) | 5,045 |
| | RVENet+ | 1.58 (1.52, 1.65) | 1.28 (1.19, 1.36) | 2.02 (1.94, 2.09) | 0.15 (0.14, 0.15) | 0.73 (0.70, 0.75) | 1.1 (0.9-1.2) | 944 |
| | EchoNet-LVH | 1.34 (1.32, 1.36) | 1.06 (1.04, 1.07) | 1.79 (1.76, 1.82) | 0.13 (0.13, 0.13) | 0.69 (0.68, 0.70) | 1.0 (0.9-1.2) | 11,681 |
| **LAIDs (mm)** | YNHHS | 3.98 (3.90, 4.05) | 3.24 (3.15, 3.33) | 5.09 (5.00, 5.19) | 0.10 (0.10, 0.10) | 0.79 (0.78, 0.80) | 4.0 (3.5-4.5) | 4,832 |
| | RVENet+ | 4.03 (3.81, 4.25) | 3.29 (3.00, 3.54) | 5.17 (4.88, 5.44) | 0.10 (0.09, 0.10) | 0.79 (0.77, 0.81) | 4.0 (3.4-4.6) | 601 |
| **LA Volume ($cm^3$)** | YNHHS | 9.41 (9.22, 9.60) | 7.89 (7.63, 8.11) | 11.86 (11.63, 12.10) | 0.16 (0.16, 0.17) | 0.78 (0.77, 0.79) | 55.2 (43.0-70.0) | 4,107 |
| | RVENet+ | 13.39 (12.88, 13.89) | 12.10 (11.11, 13.11) | 16.25 (15.63, 16.87) | 0.20 (0.20, 0.21) | 0.80 (0.78, 0.82) | 64.5 (46.2-91.3) | 845 |
| **LV ED Volume ($cm^3$)** | YNHHS | 18.97 (18.48, 19.46) | 14.26 (13.79, 14.83) | 25.97 (25.19, 26.73) | 0.17 (0.16, 0.17) | 0.86 (0.85, 0.87) | 105.1 (80.6-136.9) | 3,449 |
| | RVENet+ | 34.83 (33.12, 36.56) | 26.96 (25.30, 28.59) | 45.48 (43.30, 47.68) | 0.25 (0.24, 0.26) | 0.81 (0.78, 0.83) | 128.9 (104.0-166.1) | 944 |
| | EchoNet-Dynamic | 25.36 (24.96, 25.79) | 17.65 (17.28, 18.00) | 36.32 (35.57, 37.07) | 0.28 (0.28, 0.28) | 0.70 (0.69, 0.71) | 82.1 (62.2-108.3) | 10,032 |
| **LV ES Volume ($cm^3$)** | YNHHS | 10.41 (10.06, 10.76) | 6.94 (6.67, 7.18) | 16.06 (15.39, 16.79) | 0.21 (0.21, 0.22) | 0.90 (0.90, 0.91) | 40.1 (28.9-56.4) | 3,435 |
| | RVENet+ | 19.18 (18.09, 20.32) | 13.07 (12.26, 14.12) | 28.11 (26.23, 30.03) | 0.28 (0.28, 0.29) | 0.90 (0.88, 0.91) | 55.0 (41.9-78.0) | 944 |
| | EchoNet-Dynamic | 14.31 (14.01, 14.62) | 8.40 (8.21, 8.61) | 23.42 (22.73, 24.09) | 0.33 (0.33, 0.34) | 0.82 (0.81, 0.83) | 33.6 (23.7-49.1) | 10,032 |
| **LVIDd (mm)** | YNHHS | 3.78 (3.70, 3.85) | 3.10 (3.02, 3.16) | 4.82 (4.73, 4.90) | 0.08 (0.08, 0.08) | 0.74 (0.73, 0.75) | 4.6 (4.2-5.1) | 5,039 |
| | RVENet+ | 4.37 (4.19, 4.56) | 3.45 (3.25, 3.67) | 5.60 (5.36, 5.85) | 0.09 (0.08, 0.09) | 0.80 (0.78, 0.82) | 4.9 (4.5-5.5) | 944 |
| | EchoNet-LVH | 3.69 (3.64, 3.73) | 3.02 (2.97, 3.06) | 4.70 (4.65, 4.76) | 0.08 (0.08, 0.08) | 0.81 (0.81, 0.82) | 4.6 (4.2-5.1) | 11,713 |
| **LVIDs (mm)** | YNHHS | 3.38 (3.32, 3.45) | 2.67 (2.60, 2.74) | 4.42 (4.33, 4.52) | 0.11 (0.11, 0.11) | 0.80 (0.78, 0.81) | 3.0 (2.6-3.4) | 5,028 |
| | RVENet+ | 4.40 (4.20, 4.62) | 3.43 (3.21, 3.75) | 5.65 (5.42, 5.90) | 0.13 (0.12, 0.13) | 0.86 (0.84, 0.87) | 3.3 (2.8-3.8) | 814 |
| | EchoNet-LVH | 3.45 (3.40, 3.50) | 2.71 (2.66, 2.77) | 4.52 (4.46, 4.59) | 0.11 (0.10, 0.11) | 0.86 (0.85, 0.86) | 3.1 (2.7-3.6) | 11,268 |
| | YNHHS | 1.35 (1.32, 1.38) | 1.07 (1.03, 1.10) | 1.76 (1.72, 1.80) | 0.07 (0.06, 0.07) | 0.59 (0.58, 0.61) | 2.0 (1.9-2.2) | 3,719 |

| | | | | | | | | |
|---|---|---|---|---|---|---|---|---|
| **LVOT Diameter (mm)** | RVENet+ | 1.54 (1.47, 1.61) | 1.34 (1.25, 1.44) | 1.92 (1.84, 2.00) | 0.08 (0.07, 0.08) | 0.53 (0.47, 0.58) | 2.0 (1.9-2.2) | 614 |
| **LVPWd (mm)** | YNHHS | 1.15 (1.13, 1.17) | 0.93 (0.91, 0.96) | 1.48 (1.45, 1.50) | 0.11 (0.11, 0.12) | 0.68 (0.66, 0.69) | 1.0 (0.9-1.1) | 5,040 |
| | RVENet+ | 1.05 (1.01, 1.09) | 0.90 (0.84, 0.94) | 1.33 (1.28, 1.39) | 0.11 (0.11, 0.12) | 0.68 (0.65, 0.71) | 0.9 (0.8-1.0) | 944 |
| | EchoNet-LVH | 1.28 (1.26, 1.29) | 1.06 (1.04, 1.08) | 1.63 (1.61, 1.64) | 0.13 (0.13, 0.13) | 0.64 (0.63, 0.65) | 1.0 (0.9-1.1) | 11,672 |
| **LV Stroke Volume ($cm^3$)** | YNHHS | 11.70 (11.43, 11.98) | 9.21 (8.89, 9.41) | 15.31 (14.94, 15.69) | 0.18 (0.18, 0.18) | 0.77 (0.76, 0.78) | 62.0 (48.3-79.0) | 3,395 |
| | RVENet+ | 17.33 (16.53, 18.11) | 13.73 (12.72, 14.58) | 22.70 (21.65, 23.70) | 0.24 (0.23, 0.25) | 0.62 (0.58, 0.66) | 69.5 (56.4-84.6) | 944 |
| **RA Transverse Dimension (mm)** | YNHHS | 4.67 (4.57, 4.78) | 3.94 (3.80, 4.07) | 5.94 (5.82, 6.08) | 0.12 (0.12, 0.13) | 0.62 (0.60, 0.64) | 3.7 (3.3-4.2) | 2,897 |
| | RVENet+ | 5.15 (4.95, 5.35) | 4.43 (4.14, 4.77) | 6.35 (6.12, 6.57) | 0.13 (0.12, 0.13) | 0.57 (0.53, 0.61) | 4.0 (3.5-4.5) | 944 |
| **RVIDd (mm)** | YNHHS | 3.97 (3.87, 4.06) | 3.25 (3.13, 3.36) | 5.09 (4.98, 5.20) | 0.13 (0.12, 0.13) | 0.57 (0.54, 0.59) | 3.1 (2.7-3.5) | 3,430 |
| | RVENet+ | 3.85 (3.67, 4.03) | 3.05 (2.83, 3.27) | 5.03 (4.80, 5.25) | 0.12 (0.12, 0.13) | 0.58 (0.53, 0.62) | 3.1 (2.8-3.5) | 944 |
| **RV Systolic Pressure (mmHg)** | YNHHS | 5.98 (5.84, 6.14) | 4.60 (4.45, 4.73) | 8.11 (7.88, 8.35) | 0.20 (0.20, 0.21) | 0.68 (0.66, 0.70) | 27.5 (22.5-35.0) | 3,298 |
| | RVENet+ | 6.72 (6.30, 7.12) | 5.03 (4.65, 5.43) | 9.34 (8.67, 10.00) | 0.21 (0.20, 0.22) | 0.73 (0.70, 0.75) | 28.0 (22.0-38.0) | 689 |
| **RV S' (cm/s)** | YNHHS | 1.91 (1.86, 1.96) | 1.49 (1.44, 1.54) | 2.61 (2.52, 2.70) | 0.15 (0.14, 0.15) | 0.60 (0.58, 0.63) | 12.6 (10.9-14.7) | 3,631 |
| | RVENet+ | 1.99 (1.90, 2.08) | 1.64 (1.49, 1.76) | 2.55 (2.43, 2.68) | 0.16 (0.15, 0.17) | 0.62 (0.59, 0.66) | 13.0 (11.0-15.0) | 831 |
| **TAPSE (mm)** | YNHHS | 3.36 (3.29, 3.43) | 2.82 (2.75, 2.91) | 4.28 (4.19, 4.38) | 0.16 (0.15, 0.16) | 0.59 (0.58, 0.61) | 2.1 (1.8-2.5) | 3,734 |
| | RVENet+ | 3.78 (3.64, 3.92) | 3.26 (3.14, 3.41) | 4.68 (4.51, 4.85) | 0.18 (0.17, 0.18) | 0.72 (0.69, 0.74) | 2.1 (1.7-2.6) | 944 |
| **TV Peak Gradient (mmHg)** | YNHHS | 5.57 (5.44, 5.72) | 4.26 (4.09, 4.41) | 7.55 (7.34, 7.78) | 0.22 (0.21, 0.22) | 0.64 (0.62, 0.66) | 24.0 (19.0-30.0) | 3,515 |
| | RVENet+ | 6.48 (6.08, 6.85) | 5.10 (4.63, 5.41) | 8.86 (8.26, 9.42) | 0.26 (0.25, 0.27) | 0.68 (0.65, 0.71) | 22.0 (17.0-30.0) | 689 |

Multi-task international validation of PanEcho on continuous parameter estimation tasks. Results are presented on the internal YNHHS validation cohort and external RVENet+, EchoNet-Dynamic, and EchoNet-LVH cohorts. Values in parentheses represent bootstrapped 95% confidence intervals unless otherwise specified. Median and IQR are computed from non-missing ground truth measurements in each cohort. * = Global longitudinal strain values are presented as positive to aid readability; MAD = median absolute deviation; MAE = mean absolute error; N = number of non-missing measurements; NMAE = normalized mean absolute error; r = Pearson's correlation coefficient.

AV = aortic valve; ED = end-diastolic; ES = end-systolic; IVSd = interventricular septum thickness at diastole; IQR = interquartile range; LA = left atrium; LV = left ventricle; LAIDs = left atrial internal diameter at systole; LVIDd = left ventricular internal diameter at diastole; LVIDs = left ventricular internal diameter at systole; LVOT = left ventricular outflow tract; LVPWd = left ventricular posterior wall thickness at diastole; RA = right atrium; RV = right ventricle; RVIDd = right ventricular internal diameter at diastole; RV S' = right ventricular systolic excursion velocity; TV = tricuspid valve; YNHHS = Yale-New Haven Health System.

**eTable 5 | Detailed Performance Evaluation at the Point of Care**

| | | Dataset | AUC | AP | Sensitivity | Specificity | Brier Score | n/N |
|---|---|---|---|---|---|---|---|---|
| **AV Stenosis** | *None* | Abbreviated TTE | 0.91 (0.90, 0.93) | 0.99 (0.98, 0.99) | 0.90 (0.89, 0.91) | 0.77 (0.73, 0.80) | 0.06 (0.06, 0.07) | 2,907/3,285 |
| | | POCUS | 0.87 (0.85, 0.89) | 0.79 (0.77, 0.80) | 0.75 (0.70, 0.80) | 0.97 (0.96, 0.98) | 0.87 (0.86, 0.88) | 2,109/2,311 |
| | *Mild-Moderate* | Abbreviated TTE | 0.89 (0.88, 0.91) | 0.49 (0.44, 0.54) | 0.76 (0.72, 0.80) | 0.87 (0.86, 0.88) | 0.06 (0.06, 0.07) | 309/3,285 |
| | | POCUS | 0.85 (0.83, 0.88) | 0.76 (0.70, 0.81) | 0.77 (0.76, 0.79) | 0.20 (0.18, 0.23) | 0.32 (0.29, 0.36) | 164/2,311 |
| | *Severe* | Abbreviated TTE | 0.96 (0.94, 0.97) | 0.42 (0.33, 0.53) | 0.83 (0.75, 0.90) | 0.94 (0.93, 0.95) | 0.02 (0.01, 0.02) | 69/3,285 |
| | | POCUS | 0.86 (0.81, 0.92) | 0.55 (0.41, 0.68) | 0.96 (0.95, 0.96) | 0.17 (0.12, 0.23) | 0.26 (0.19, 0.34) | 38/2,311 |
| | *Mean* | Abbreviated TTE | 0.92 (0.91, 0.93) | 0.63 (0.60, 0.67) | 0.83 (0.80, 0.86) | 0.86 (0.85, 0.87) | 0.05 (0.04, 0.05) | N/A |
| | | POCUS | 0.86 (0.84, 0.88) | 0.70 (0.65, 0.75) | 0.83 (0.81, 0.84) | 0.45 (0.43, 0.47) | 0.48 (0.46, 0.51) | N/A |
| **AV Structure** | *Any* | Abbreviated TTE | 0.77 (0.70, 0.84) | 1.00 (0.99, 1.00) | 0.75 (0.74, 0.76) | 0.68 (0.56, 0.79) | 0.01 (0.01, 0.01) | 37/3,219 |
| | | POCUS | 0.70 (0.56, 0.85) | 0.95 (0.94, 0.95) | 0.30 (0.00, 0.56) | 1.00 (1.00, 1.00) | 0.97 (0.97, 0.97) | 10/2,567 |
| **Increased LA Size** | *Mild* | Abbreviated TTE | 0.74 (0.72, 0.76) | 0.36 (0.33, 0.39) | 0.68 (0.65, 0.71) | 0.67 (0.66, 0.68) | 0.12 (0.11, 0.13) | 570/3,608 |
| | | POCUS | 0.63 (0.61, 0.65) | 0.50 (0.46, 0.55) | 0.67 (0.65, 0.68) | 0.17 (0.16, 0.19) | 0.26 (0.23, 0.28) | 403/3,310 |
| | *≥ Moderate* | Abbreviated TTE | 0.87 (0.86, 0.88) | 0.56 (0.53, 0.60) | 0.74 (0.72, 0.77) | 0.82 (0.81, 0.83) | 0.10 (0.09, 0.10) | 598/3,608 |
| | | POCUS | 0.78 (0.77, 0.80) | 0.67 (0.64, 0.69) | 0.74 (0.72, 0.75) | 0.42 (0.40, 0.45) | 0.52 (0.49, 0.54) | 741/3,310 |
| | *None* | Abbreviated TTE | 0.82 (0.81, 0.83) | 0.91 (0.90, 0.91) | 0.72 (0.71, 0.74) | 0.76 (0.74, 0.78) | 0.16 (0.15, 0.16) | 2,440/3,608 |
| | | POCUS | 0.72 (0.71, 0.74) | 0.62 (0.60, 0.64) | 0.72 (0.70, 0.74) | 0.81 (0.79, 0.82) | 0.70 (0.69, 0.72) | 2,166/3,310 |
| | *Mean* | Abbreviated TTE | 0.81 (0.80, 0.82) | 0.61 (0.59, 0.63) | 0.72 (0.70, 0.73) | 0.75 (0.74, 0.76) | 0.13 (0.12, 0.13) | N/A |
| | | POCUS | 0.71 (0.70, 0.73) | 0.60 (0.58, 0.61) | 0.71 (0.70, 0.72) | 0.47 (0.46, 0.48) | 0.49 (0.48, 0.50) | N/A |
| **LV Diastolic Dysfunction** | *Mild* | Abbreviated TTE | 0.79 (0.78, 0.80) | 0.63 (0.61, 0.66) | 0.61 (0.58, 0.64) | 0.79 (0.77, 0.80) | 0.17 (0.16, 0.17) | 892/2,825 |
| | | POCUS | 0.65 (0.63, 0.68) | 0.73 (0.69, 0.76) | 0.48 (0.45, 0.50) | 0.34 (0.31, 0.36) | 0.46 (0.43, 0.49) | 473/1,769 |
| | *≥ Moderate* | Abbreviated TTE | 0.91 (0.89, 0.92) | 0.51 (0.46, 0.57) | 0.85 (0.81, 0.88) | 0.83 (0.82, 0.84) | 0.06 (0.05, 0.06) | 247/2,825 |
| | | POCUS | 0.83 (0.82, 0.85) | 0.79 (0.75, 0.82) | 0.74 (0.72, 0.76) | 0.48 (0.45, 0.51) | 0.60 (0.57, 0.62) | 416/1,769 |
| | *None* | Abbreviated TTE | 0.84 (0.83, 0.85) | 0.89 (0.88, 0.90) | 0.80 (0.78, 0.81) | 0.70 (0.68, 0.72) | 0.16 (0.16, 0.17) | 1,686/2,825 |
| | | POCUS | 0.80 (0.79, 0.82) | 0.59 (0.56, 0.62) | 0.85 (0.83, 0.87) | 0.79 (0.77, 0.82) | 0.68 (0.65, 0.70) | 880/1,769 |

| | | | | | | | | |
|---|---|---|---|---|---|---|---|---|
| | *Mean* | Abbreviated TTE | 0.85 (0.84, 0.86) | 0.68 (0.66, 0.70) | 0.75 (0.73, 0.77) | 0.77 (0.76, 0.78) | 0.13 (0.12, 0.13) | N/A |
| | | POCUS | 0.76 (0.75, 0.78) | 0.70 (0.68, 0.72) | 0.69 (0.68, 0.70) | 0.54 (0.52, 0.55) | 0.58 (0.56, 0.59) | N/A |
| **Elevated LVOT PG** | *≥ 20 mmHg* | Abbreviated TTE | 0.94 (0.87, 0.99) | 0.54 (0.37, 0.71) | 0.73 (0.56, 0.88) | 0.96 (0.95, 0.96) | 0.01 (0.00, 0.01) | 22/3,096 |
| | | POCUS | N/A | N/A | N/A | N/A | N/A | 2/2,433 |
| **Increased LV Size** | *Mild* | Abbreviated TTE | 0.83 (0.81, 0.85) | 0.30 (0.26, 0.35) | 0.69 (0.64, 0.74) | 0.80 (0.79, 0.81) | 0.07 (0.06, 0.07) | 253/2,981 |
| | | POCUS | 0.64 (0.62, 0.67) | 0.50 (0.46, 0.55) | 0.68 (0.67, 0.70) | 0.20 (0.17, 0.22) | 0.28 (0.26, 0.31) | 351/2,632 |
| | *≥ Moderate* | Abbreviated TTE | 0.94 (0.92, 0.95) | 0.63 (0.58, 0.69) | 0.76 (0.71, 0.81) | 0.92 (0.91, 0.92) | 0.04 (0.03, 0.04) | 201/2,981 |
| | | POCUS | 0.89 (0.87, 0.90) | 0.72 (0.69, 0.75) | 0.89 (0.88, 0.90) | 0.64 (0.61, 0.67) | 0.68 (0.65, 0.70) | 547/2,632 |
| | *None* | Abbreviated TTE | 0.90 (0.89, 0.91) | 0.98 (0.97, 0.98) | 0.88 (0.87, 0.89) | 0.72 (0.69, 0.76) | 0.08 (0.07, 0.09) | 2,527/2,981 |
| | | POCUS | 0.85 (0.84, 0.86) | 0.88 (0.86, 0.89) | 0.67 (0.65, 0.70) | 0.84 (0.82, 0.85) | 0.86 (0.85, 0.87) | 1,734/2,632 |
| | *Mean* | Abbreviated TTE | 0.89 (0.88, 0.90) | 0.64 (0.62, 0.66) | 0.78 (0.75, 0.80) | 0.81 (0.80, 0.82) | 0.06 (0.06, 0.07) | N/A |
| | | POCUS | 0.79 (0.78, 0.81) | 0.70 (0.68, 0.72) | 0.75 (0.74, 0.76) | 0.56 (0.55, 0.57) | 0.61 (0.59, 0.62) | N/A |
| **LV Systolic Dysfunction** | *Mild* | Abbreviated TTE | 0.91 (0.89, 0.92) | 0.36 (0.32, 0.42) | 0.83 (0.79, 0.87) | 0.85 (0.84, 0.86) | 0.05 (0.04, 0.05) | 223/3,440 |
| | | POCUS | 0.70 (0.68, 0.72) | 0.71 (0.68, 0.75) | 0.58 (0.57, 0.60) | 0.21 (0.20, 0.23) | 0.33 (0.31, 0.35) | 430/3,114 |
| | *≥ Moderate* | Abbreviated TTE | 0.98 (0.97, 0.98) | 0.81 (0.77, 0.85) | 0.86 (0.82, 0.90) | 0.95 (0.95, 0.96) | 0.02 (0.02, 0.03) | 212/3,440 |
| | | POCUS | 0.93 (0.92, 0.94) | 0.89 (0.87, 0.91) | 0.80 (0.79, 0.82) | 0.59 (0.57, 0.62) | 0.71 (0.69, 0.73) | 755/3,114 |
| | *None* | Abbreviated TTE | 0.96 (0.95, 0.97) | 0.99 (0.99, 0.99) | 0.91 (0.90, 0.92) | 0.86 (0.83, 0.89) | 0.05 (0.04, 0.05) | 3,005/3,440 |
| | | POCUS | 0.89 (0.88, 0.90) | 0.71 (0.69, 0.73) | 0.87 (0.85, 0.88) | 0.90 (0.88, 0.91) | 0.79 (0.78, 0.80) | 1,929/3,114 |
| | *Mean* | Abbreviated TTE | 0.95 (0.94, 0.95) | 0.72 (0.70, 0.74) | 0.87 (0.85, 0.88) | 0.89 (0.88, 0.90) | 0.04 (0.04, 0.04) | N/A |
| | | POCUS | 0.84 (0.83, 0.85) | 0.77 (0.76, 0.79) | 0.75 (0.74, 0.76) | 0.57 (0.56, 0.58) | 0.61 (0.60, 0.62) | N/A |
| **LV Wall Motion Abnormalities** | *Any* | Abbreviated TTE | 0.86 (0.84, 0.88) | 0.72 (0.67, 0.76) | 0.73 (0.68, 0.78) | 0.83 (0.80, 0.85) | 0.11 (0.10, 0.12) | 249/1,096 |
| | | POCUS | 0.77 (0.75, 0.79) | 0.81 (0.78, 0.83) | 0.53 (0.50, 0.56) | 0.58 (0.55, 0.61) | 0.68 (0.65, 0.70) | 630/1,415 |
| **Increased LV Wall Thickness** | *Any* | Abbreviated TTE | 0.85 (0.84, 0.86) | 0.89 (0.88, 0.90) | 0.82 (0.80, 0.83) | 0.70 (0.68, 0.72) | 0.15 (0.15, 0.16) | 1,465/3,434 |
| | | POCUS | 0.75 (0.73, 0.76) | 0.37 (0.35, 0.39) | 0.90 (0.89, 0.92) | 0.83 (0.80, 0.85) | 0.51 (0.49, 0.53) | 1,206/2,735 |
| | *≥ Moderate* | Abbreviated TTE | 0.93 (0.91, 0.94) | 0.99 (0.99, 0.99) | 0.90 (0.89, 0.91) | 0.79 (0.74, 0.83) | 0.04 (0.04, 0.05) | 250/3,355 |
| | | POCUS | 0.85 (0.83, 0.87) | 0.70 (0.68, 0.71) | 0.83 (0.79, 0.86) | 0.97 (0.97, 0.98) | 0.81 (0.80, 0.82) | 258/2,710 |

| | | | | | | | | |
|---|---|---|---|---|---|---|---|---|
| **MV Stenosis** | *Any* | Abbreviated TTE | 0.94 (0.92, 0.96) | 1.00 (1.00, 1.00) | 0.97 (0.96, 0.97) | 0.69 (0.59, 0.78) | 0.02 (0.01, 0.02) | 67/2,915 |
| | | POCUS | 0.92 (0.88, 0.95) | 0.95 (0.95, 0.96) | 0.63 (0.54, 0.72) | 0.99 (0.98, 0.99) | 0.97 (0.97, 0.97) | 71/2,062 |
| **Pericardial Effusion** | *Any* | Abbreviated TTE | 0.91 (0.82, 0.97) | 0.99 (0.98, 1.00) | 0.91 (0.89, 0.93) | 0.76 (0.60, 0.91) | 0.02 (0.02, 0.03) | 21/619 |
| | | POCUS | 0.86 (0.84, 0.88) | 0.48 (0.46, 0.50) | 0.93 (0.90, 0.95) | 0.98 (0.97, 0.99) | 0.64 (0.63, 0.66) | 305/2,796 |
| **Elevated RA Pressure** | *≥ 8 mmHg* | Abbreviated TTE | 0.80 (0.78, 0.82) | 0.48 (0.44, 0.53) | 0.62 (0.58, 0.66) | 0.82 (0.81, 0.83) | 0.10 (0.09, 0.11) | 485/3,289 |
| | | POCUS | 0.75 (0.74, 0.77) | 0.96 (0.95, 0.97) | 0.22 (0.20, 0.23) | 0.48 (0.47, 0.50) | 0.64 (0.63, 0.66) | 1,200/2,775 |
| **Increased RA Size** | *Any* | Abbreviated TTE | 0.75 (0.72, 0.77) | 0.87 (0.84, 0.89) | 0.70 (0.68, 0.73) | 0.70 (0.66, 0.74) | 0.18 (0.17, 0.19) | 374/1,267 |
| | | POCUS | 0.74 (0.72, 0.77) | 0.36 (0.34, 0.39) | 0.89 (0.87, 0.91) | 0.82 (0.79, 0.86) | 0.50 (0.47, 0.53) | 541/1,310 |
| **Increased RV Size** | *Mild* | Abbreviated TTE | 0.76 (0.74, 0.78) | 0.24 (0.21, 0.27) | 0.57 (0.52, 0.61) | 0.77 (0.76, 0.79) | 0.09 (0.08, 0.10) | 359/3,360 |
| | | POCUS | 0.67 (0.64, 0.69) | 0.79 (0.76, 0.82) | 0.46 (0.44, 0.47) | 0.21 (0.19, 0.22) | 0.33 (0.31, 0.35) | 427/2,793 |
| | *≥ Moderate* | Abbreviated TTE | 0.88 (0.86, 0.90) | 0.27 (0.22, 0.33) | 0.69 (0.63, 0.75) | 0.86 (0.85, 0.87) | 0.04 (0.03, 0.04) | 158/3,360 |
| | | POCUS | 0.87 (0.86, 0.89) | 0.97 (0.96, 0.98) | 0.38 (0.37, 0.40) | 0.21 (0.20, 0.23) | 0.35 (0.33, 0.37) | 411/2,793 |
| | *None* | Abbreviated TTE | 0.82 (0.80, 0.84) | 0.96 (0.95, 0.96) | 0.84 (0.82, 0.85) | 0.61 (0.58, 0.65) | 0.11 (0.10, 0.11) | 2,843/3,360 |
| | | POCUS | 0.82 (0.80, 0.83) | 0.48 (0.46, 0.49) | 0.89 (0.88, 0.91) | 0.91 (0.90, 0.92) | 0.63 (0.61, 0.64) | 1,955/2,793 |
| | *Mean* | Abbreviated TTE | 0.82 (0.81, 0.83) | 0.49 (0.47, 0.51) | 0.70 (0.67, 0.72) | 0.75 (0.74, 0.76) | 0.08 (0.07, 0.08) | N/A |
| | | POCUS | 0.79 (0.77, 0.80) | 0.75 (0.73, 0.76) | 0.58 (0.57, 0.59) | 0.44 (0.44, 0.45) | 0.43 (0.42, 0.45) | N/A |
| **RV Systolic Dysfunction** | *Any* | Abbreviated TTE | 0.92 (0.90, 0.93) | 0.99 (0.99, 0.99) | 0.90 (0.89, 0.91) | 0.74 (0.69, 0.79) | 0.05 (0.04, 0.05) | 263/3,347 |
| | | POCUS | 0.85 (0.84, 0.86) | 0.71 (0.69, 0.72) | 0.81 (0.79, 0.83) | 0.90 (0.89, 0.91) | 0.79 (0.78, 0.80) | 819/2,813 |

Multi-task validation of PanEcho on diagnostic classification tasks. Results are presented on the internal YNHHS validation cohort and external RVENet+, EchoNet-Dynamic, and EchoNet-LVH cohorts. Values in parentheses represent bootstrapped 95% confidence intervals. For multi-class classification tasks, metrics are presented for each individual class, followed by the "Mean" overall performance across classes. n = number of studies meeting the label definition; N = number of non-missing labels; * = moderate or higher; ^ = severe.
AP = average precision; AUC = area under the receiver operator characteristic curve; AV = aortic valve; LA = left atrium; LV = left ventricle; LVOT = left ventricular outflow tract; PG = pressure gradient; RA = right atrium; RV = right ventricle; TV = tricuspid valve; YNHHS = Yale-New Haven Health System.

#### eFigure 1 | Bland-Altman Analysis for Key Echocardiographic Measurements

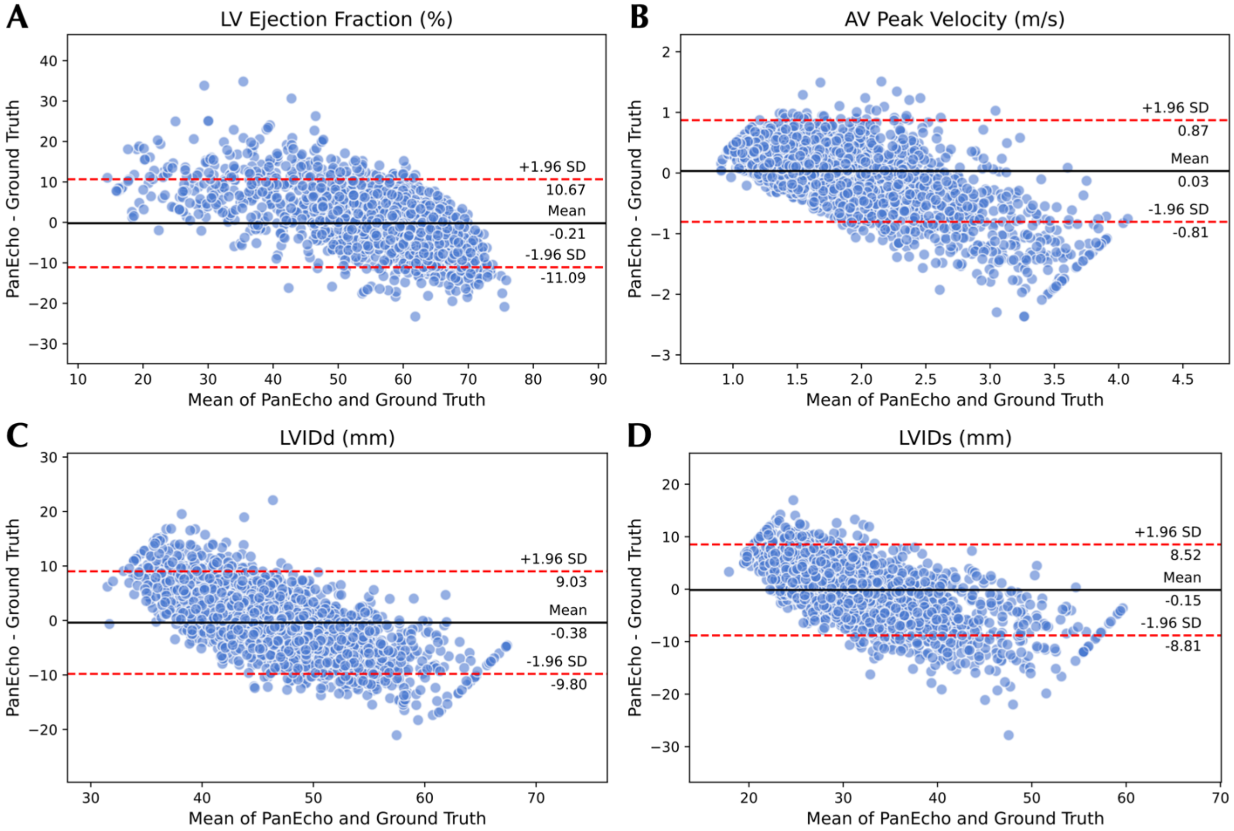


Bland-Altman plots for LV ejection fraction (A), AV peak velocity (B), LVIDd (C), and LVIDs (D). Analysis compares the automated PanEcho estimation vs. human expert ground truth measurement in the internal YNHHS validation set. Limits of agreement are determined by 1.96 times the standard deviation of differences.

AV = aortic valve; LV = left ventricle; LVIDd = left ventricular internal diameter at diastole; LVIDs = left ventricular internal diameter at systole; YNHHS = Yale-New Haven Health System.

#### eFigure 2 | Task-specific View Relevance Analysis

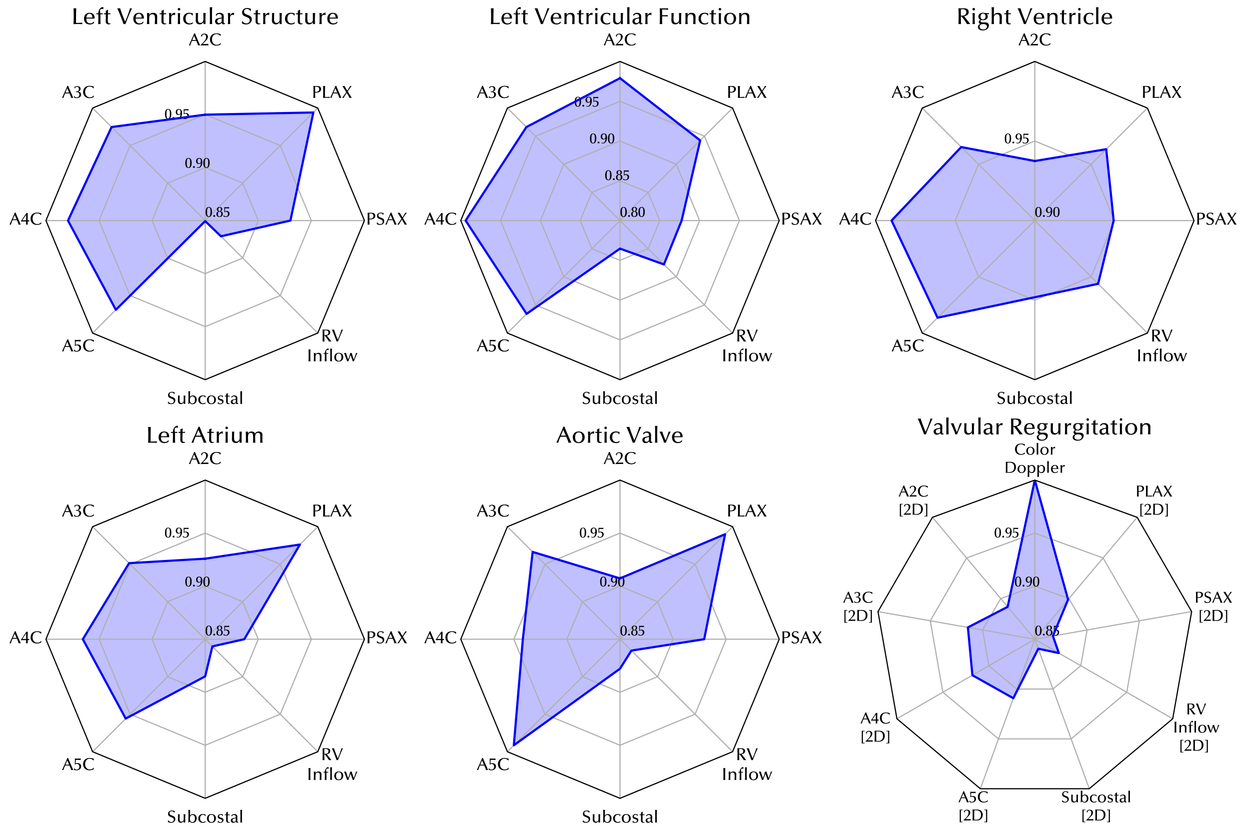


Radar plots depicting the relative importance of each echocardiographic view for a given aspect of cardiovascular diagnosis. For each task, a normalized view relevance score is computed, where 1 indicates the most relevant view; this score reflects the fraction of the maximum performance metric (AUC for classification tasks and MAE for regression tasks) achieved on a given task when only using videos from an individual view. Presented view relevance scores are then averaged over all tasks falling under a given category in each subplot. Analysis is performed on the internal YNHHS validation set using up to three videos from a given echocardiographic view per study.
A2C = apical 2-chamber; A3C = apical 3-chamber; A4C = apical 4-chamber; A5C = apical 5-chamber; AUC = area under the receiver operator characteristic curve; PLAX = parasternal long axis; PSAX = parasternal short axis; YNHHS = Yale-New Haven Health System.

#### eFigure 3 | Group Fairness Analysis

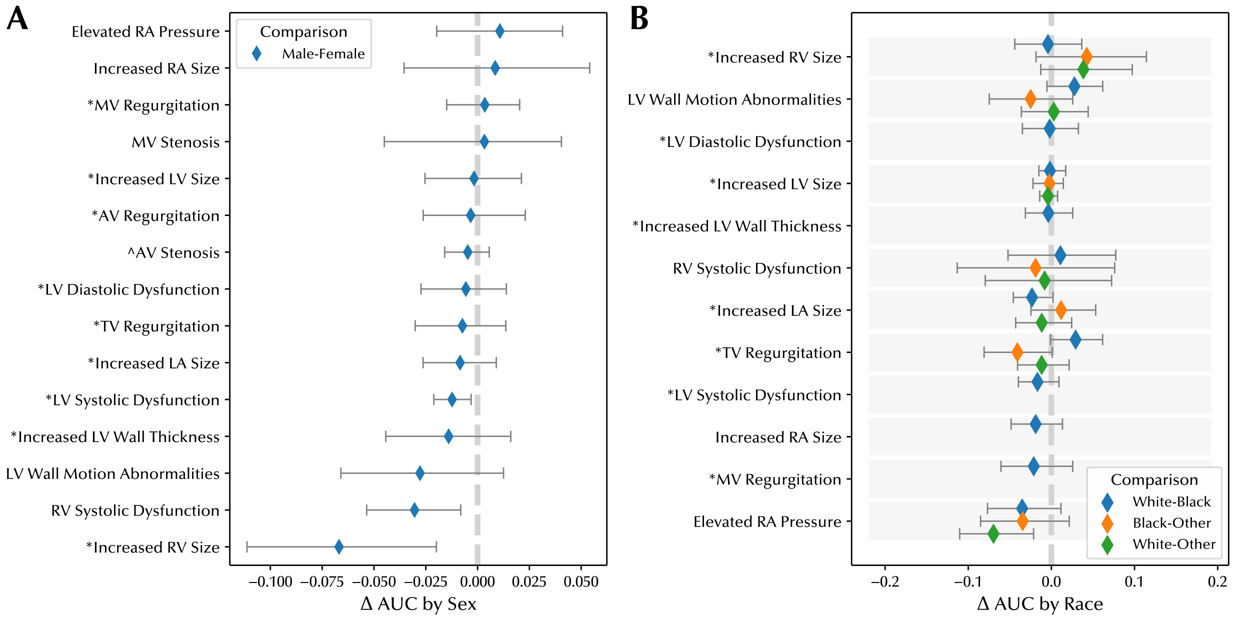


Fairness analysis depicting the difference in AUC between different protect subgroups by sex (A) and race (B). Groupwise comparisons were excluded if the subgroup had <30 positive examples of a particular task to ensure reliable sufficient sample size. Analysis was performed in the internal YNHHS validation set. Error bars and values in parentheses represent bootstrapped 95% confidence intervals. Grey dashed line represents parity between subgroups. * = moderate or higher, ^ = severe.
AUC = area under the receiver operator characteristic curve; AV = aortic valve; LA = left atrium; LV = left ventricle; LVOT = left ventricular outflow tract; PG = pressure gradient; RA = right atrium; RV = right ventricle; TV = tricuspid valve; YNHHS = Yale-New Haven Health System.

**eFigure 4 | Image Quality Analysis**

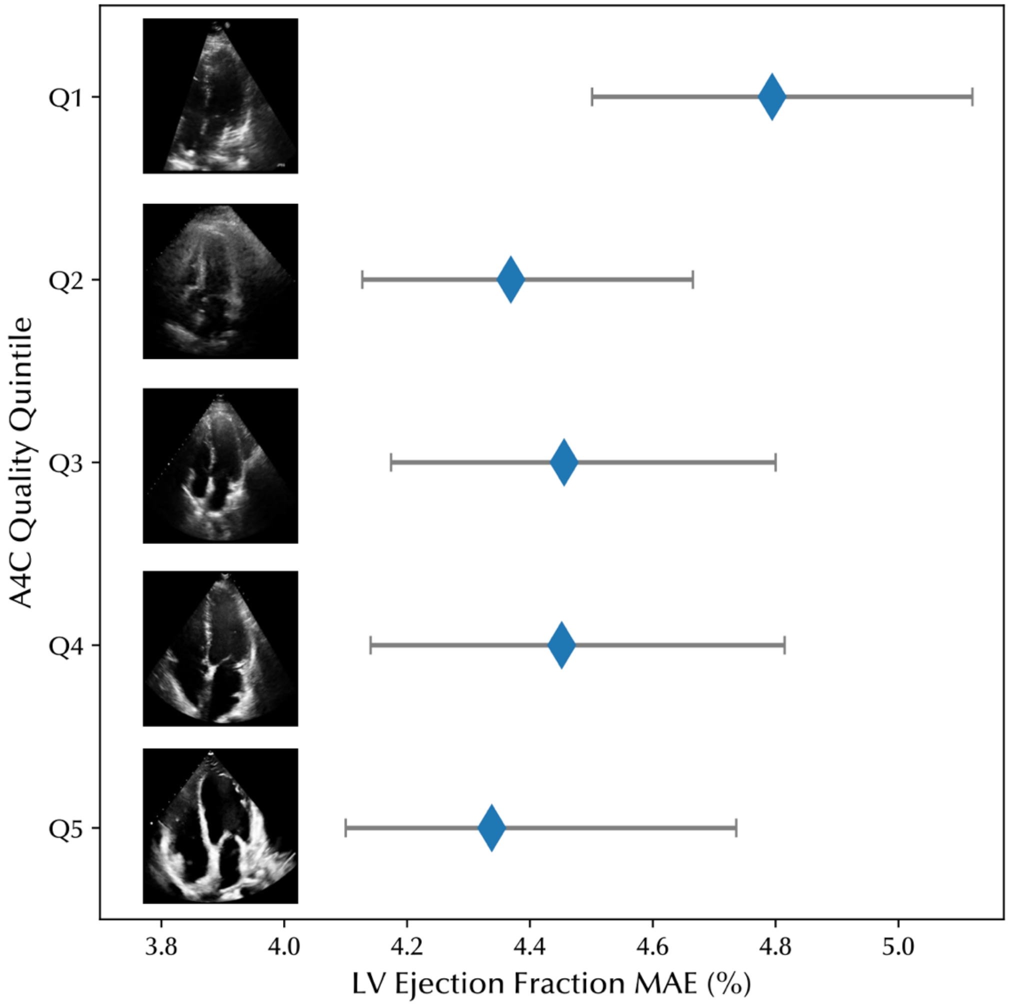


LV ejection fraction estimation performance, measured by MAE, across equal-sized "bins" of image quality. Only A4C videos from the internal YNHHS validation cohort were used for this analysis. Each bin represents a bin of image quality, defined by the quintile of A4C view confidence as predicted by the pretrained view classifier. A representative example frame from each quintile is presented alongside each row. Error bars represent bootstrapped 95% confidence intervals.
A4C = apical 4-chamber; LV = left ventricle; MAE = mean absolute error.

**eReferences**


1. Holste G, Oikonomou EK, Mortazavi BJ, et al. Severe aortic stenosis detection by deep learning applied to echocardiography. *Eur Heart J*. Published online August 23, 2023. doi:10.1093/eurheartj/ehad456

2. Zhang J, Gajjala S, Agrawal P, et al. Fully Automated Echocardiogram Interpretation in Clinical Practice. *Circulation*. 2018;138(16):1623-1635.

3. Liu Z, Mao H, Wu C, Feichtenhofer C, Darrell T, Xie S. A ConvNet for the 2020s. *Proc IEEE Comput Soc Conf Comput Vis Pattern Recognit*. Published online January 10, 2022:11966-11976.

4. Kingma DP, Ba J. Adam: A Method for Stochastic Optimization. *arXiv [csLG]*. Published online December 22, 2014. http://arxiv.org/abs/1412.6980

5. Deng J, Dong W, Socher R, Li LJ, Li K, Fei-Fei L. ImageNet: A large-scale hierarchical image database. In: *2009 IEEE Conference on Computer Vision and Pattern Recognition*. IEEE; 2009:248-255.

6. Vaswani A, Shazeer NM, Parmar N, et al. Attention is All you Need. *Adv Neural Inf Process Syst*. Published online June 12, 2017:5998-6008.

7. Srivastava N, Hinton G, Krizhevsky A, Sutskever I, Salakhutdinov R. Dropout: A Simple Way to Prevent Neural Networks from Overfitting. *J Mach Learn Res*. 2014;15(56):1929-1958.

8. Paszke, Gross, Massa, Lerer. Pytorch: An imperative style, high-performance deep learning library. *Adv Neural Inf Process Syst*. Published online 2019. https://proceedings.neurips.cc/paper/2019/hash/bdbca288fee7f92f2bfa9f7012727740-Abstract.html

9. Zoghbi WA, Adams D, Bonow RO, et al. Recommendations for noninvasive evaluation of native valvular regurgitation: A report from the American society of echocardiography developed in collaboration with the society for cardiovascular magnetic resonance. *J Am Soc Echocardiogr*. 2017;30(4):303-371.

10. Baumgartner H Chair, Hung J Co Chair, Bermejo J, et al. Recommendations on the echocardiographic assessment of aortic valve stenosis: a focused update from the European Association of Cardiovascular Imaging and the American Society of Echocardiography. *Eur Heart J Cardiovasc Imaging*. 2017;18(3):254-275.

11. Baumgartner H, Hung J, Bermejo J, et al. Echocardiographic assessment of valve stenosis: EAE/ASE recommendations for clinical practice. *J Am Soc Echocardiogr*. 2009;22(1):1-23; quiz 101-102.

12. Nagueh SF, Smiseth OA, Appleton CP, et al. Recommendations for the evaluation of left ventricular diastolic function by echocardiography: An update from the American society of echocardiography and the European association of cardiovascular imaging. *J Am Soc Echocardiogr*. 2016;29(4):277-314.
